# Tau accumulation and its spatial progression across the Alzheimer’s disease spectrum

**DOI:** 10.1101/2023.06.02.23290880

**Authors:** Frédéric St-Onge, Marianne Chapleau, John CS Breitner, Sylvia Villeneuve, Alexa Pichet Binette, the Alzheimer’s Disease Neuroimaging Initiative

## Abstract

The spread of tau abnormality in sporadic Alzheimer’s disease is believed typically to follow neuropathologically defined Braak staging. Recent *in-vivo* positron emission tomography (PET) evidence challenges this belief, however, as spreading patterns for tau appear heterogenous among individuals with varying clinical expression of Alzheimer’s disease. We therefore sought better understanding of the spatial distribution of tau in the preclinical and clinical phases of sporadic Alzheimer’s disease and its association with cognitive decline.

Longitudinal tau-PET data (1,370 scans) from 832 participants (463 cognitively unimpaired, 277 with mild cognitive impairment (MCI) and 92 with Alzheimer’s disease dementia) were obtained from the Alzheimer’s Disease Neuroimaging Initiative. Among these, we defined thresholds of abnormal tau deposition in 70 brain regions from the Desikan atlas, and for each group of regions characteristic of Braak staging. We summed each scan’s number of regions with abnormal tau deposition to form a spatial extent index. We then examined patterns of tau pathology cross-sectionally and longitudinally and assessed their heterogeneity. Finally, we compared our spatial extent index of tau uptake with a temporal meta region of interest—a commonly used proxy of tau burden—assessing their association with cognitive scores and clinical progression.

More than 80% of amyloid-beta positive participants across diagnostic groups followed typical Braak staging, both cross-sectionally and longitudinally. Within each Braak stage, however, the pattern of abnormality demonstrated significant heterogeneity such that overlap of abnormal regions across participants averaged less than 50%. The annual rate of change in number of abnormal tau-PET regions was similar among individuals without cognitive impairment and those with Alzheimer’s disease dementia. Spread of disease progressed more rapidly, however, among participants with MCI. The latter’s change on our spatial extent measure amounted to 2.5 newly abnormal regions per year, as contrasted with 1 region/year among the other groups. Comparing the association of tau pathology and cognitive performance in MCI and Alzheimer’s disease dementia, our spatial extent index was superior to the temporal meta-ROI for measures of executive function.

Thus, while participants broadly followed Braak stages, significant individual regional heterogeneity of tau binding was observed at each clinical stage. Progression of spatial extent of tau pathology appears to be fastest in persons with MCI. Exploring the spatial distribution of tau deposits throughout the entire brain may uncover further pathological variations and their correlation with impairments in cognitive functions beyond memory.

## Introduction

The first positron emission tomography (PET) tracers of tau pathology were developed almost a decade ago.^1^ These tracers have advanced our understanding of the role of tau pathology in aging and Alzheimer’s disease.^2–5^ However, several questions remain, including the spatial progression of the disease across the whole brain. Our principal aim was to provide a comprehensive view of the prevalence and the clinical relevance of cross-sectional and longitudinal tau-PET binding in late onset sporadic Alzheimer’s disease. Using data from the Alzheimer’s disease neuroimaging initiative (ADNI) we here report the prevalence of abnormal tau PET binding in individuals classified as being cognitively unimpaired [CU] or having mild cognitive impairment [MCI] or Alzheimer’s disease dementia. We also report the amount and the spatial extent of tau abnormality across these clinical groups both cross-sectionally and over time. Finally, we describe their association with cognitive impairment.

The progression of tau pathology in the brain is generally believed to follow a stereotypical pattern approximating the Braak stages defined post-mortem, where tau starts accumulating in medial temporal regions (Braak I-II) before spreading to limbic regions (Braak III-IV) and finally to the whole cortical mantle (Braak V-VI).^6^ Many PET studies have confirmed this pattern in-vivo,^5, 7, 8^ and studies investigating associations between tau and clinical variables usually average tau from a predefined set of temporal regions (i.e., a temporal meta-region of interest or ROI) to approximate the early stages of tau spreading.^9–11^

Reports in recent years have highlighted the limitations of this homogenous approach, however as tau progression patterns can differ across individuals^12,13^ and between different disease variants.^14–16^ These inter-individual differences would seem important to track longitudinal changes, and it has been suggested that tau accumulation and spreading are better captured when using individualized ROIs.^12,17^ Inter-individual differences in tau pathology may become particularly critical when tracking clinical progression. Evidence thus far highlights that tau, rather than amyloid-beta (Aβ) alone, is a reliable indicator of future clinical progression,^11,18^ and is well associated with cognitive change in early stages of Alzheimer’s disease.^19–, 23^ Therefore, if tau patterns and their progression are indeed heterogenous, it appears likely that tracking tau with a single set of regions across participants may misrepresent a significant portion of them.

Leveraging 1,370 tau PET scan visits from 832 ADNI participants across the Alzheimer’s disease spectrum, we characterized the spatial extent of tau pathology across the whole brain (70 brain regions) both cross-sectionally and longitudinally. We summarized these measures by developing a novel spatial extent index. This index accounts for individual differences in tau-PET patterns by evaluating the extent of tau pathology for any single individual across the whole brain. We then evaluated how the spatial extent index related to performance on different cognitive domains. We compared this approach with more traditional measures of Braak staging and tau-PET uptake in a temporal meta-region of interest (ROI).^9^ We hypothesized that a region-specific analysis of tau-PET abnormality would offer a more useful measure of cognitive impairment than other approaches that rely on tracer uptake in one set of regions across all individuals.

In summary, our study revealed that although participants exhibited an accumulation of abnormal tau levels that aligned with the predefined Braak stages, there was significant heterogeneity among individuals regarding the specific regions affected. Additionally, while the rate of local tau accumulation, measured by the average SUVR change within regions of interest (ROIs), was similar between individuals with MCI and dementia, the spreading of tau pathology (number of regions progressing from tau-negative to tau-positive) was more rapid in those with MCI. Furthermore, when it came to capturing executive function cognitive deficits, the spatial extent index performed slightly better than the temporal meta-ROI. However, both measures were equally associated with memory performance.

## Materials and methods

### Participants

We used data from ADNI, a multi-site study launched in 2003 as a public-private partnership. The primary goal of ADNI has been to test whether serial MRI, PET, other biological markers, and clinical and neuropsychological assessment can be combined to measure the progression of MCI and early Alzheimer’s disease. For up-to-date information, see www.adni-info.org.

We conducted the analyses using ADNI longitudinal data available in May 2022. We included participants who had at least one available tau (flortaucipir) and one Aβ (florbetapir or florbetaben) PET scan, and who had an available diagnostic assessment within two years from the tau scan in ADNI3.

### PET acquisition and processing

We used fully preprocessed data from the ADNI consortium. Details on PET acquisition and preprocessing procedures can be found elsewhere (http://adni.loni.usc.edu/methods/documents/). Briefly, for tau-PET, the flortaucipir tracer ([^18^F] AV-1451) was used and images were acquired 75-105 minutes post-injection. For Aβ-PET, florbetapir or florbetaben were used, and images were acquired 50-70-and 90-110-minutes post-injection, respectively. Briefly, PET images were realigned, averaged, resliced to 1.5mm^3^ and smoothed to a resolution of 8mm^3^ full width at half-maximum. Then, the closest T1-weighted MRI available for a participant was processed and segmented using FreeSurfer 7.1.1, and co-registered to the PET scan using SPM. SUVRs were extracted from each cortical region of the Desikan atlas.^24^ The inferior cerebellum was used as the reference region for flortaucipir, and the whole cerebellum was the reference region for Aβ-PET. As suggested by the ADNI PET core group, we divided the SUVR values provided by ADNI by the SUVR values in the reference region for each tracer.

Aβ-PET positivity status was determined according to the cutoff derived from the ADNI PET core based on a neocortial composite region: participants exceeding 1.11 SUVR for florbetapir or 1.08 SUVR for florbetaben were considered positive. We also converted the SUVR values into centiloid units for supplementary analyses, following established formulas from the ADNI PET core.^25^

### Regional tau-PET and other measures of interest

Our main interest was to study the patterns of elevated regional tau-PET uptake across the brain at the individual level. For this aim, we derived an SUVR cutoff for each brain region of interest using Gaussian-mixture modeling (GMM) on the entire cross-sectional sample of ADNI participants. This procedure is illustrated in Figure 1. We fitted a two-component GMM for each region and used the SUVR closest to the 50% probability of belonging to the abnormal (high values) distribution as the regional cutoff, as done previously.^26, 27^ The brain regions of interest were the 34 bilateral cortical regions of the Desikan atlas^24^ and the amygdalae. We then binarized the tau SUVR from each region, and values at or exceeding the cutoff were coded as one and a score lower than the cutoff as zero. From there, we derived our main measure of interest: the spatial extent index, which is the sum of regions exceeding the regional thresholds for a given participant. Regional thresholds for each region are provided in Supplementary Table 1.

**Figure 1.**
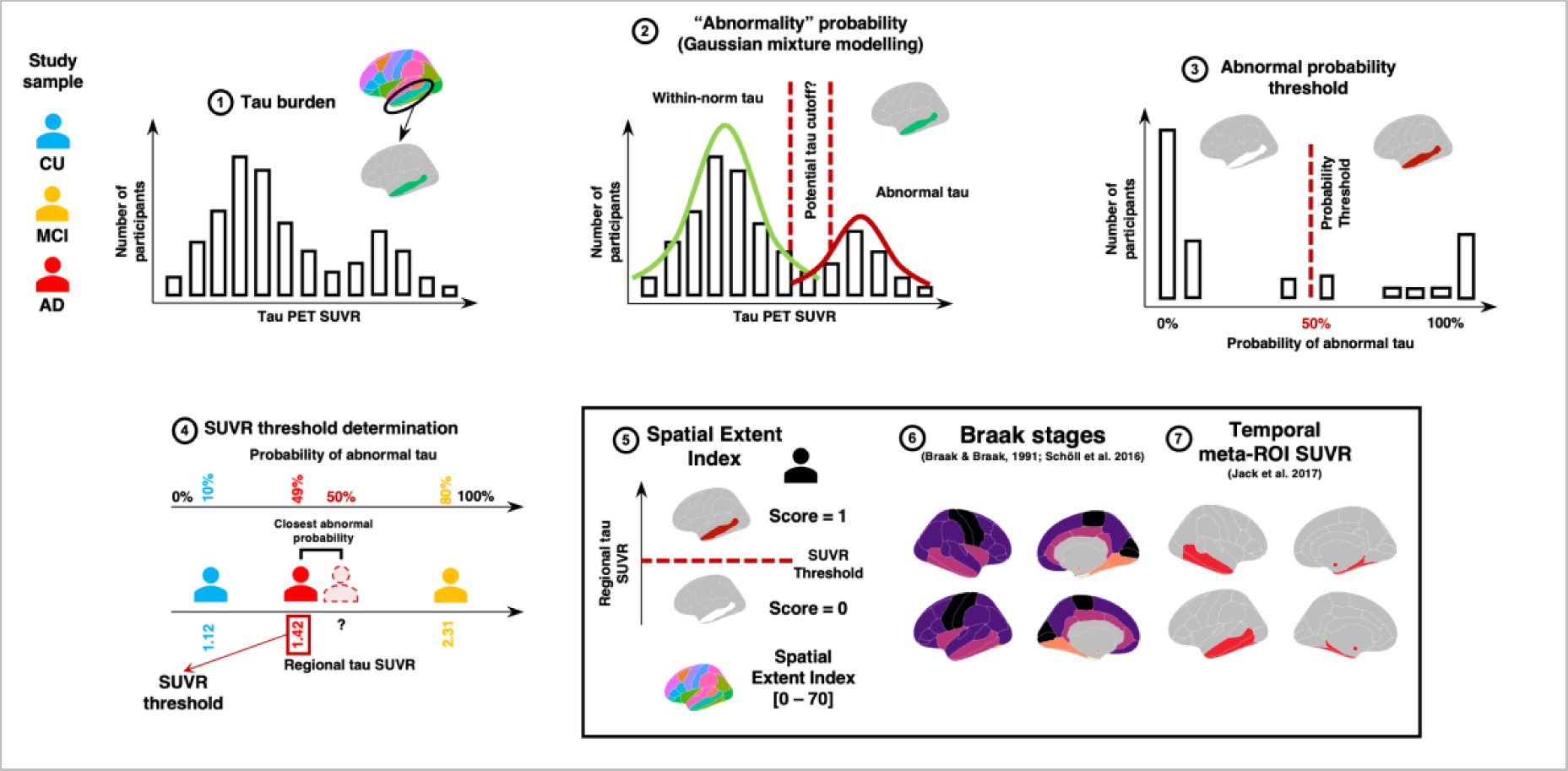
Spatial extent methodology. For each cortical region of the Desikan atlas and the bilateral amygdalae, we extract the standardized uptake value ratio (SUVR) of our participants (**1**). Then, a two-component gaussian mixture modelling technique is applied to the SUVR values in each region (**2-3**). The second distribution is considered to reflect abnormally high SUVR tau values. We extract the probability that each participant belongs to the “abnormal” distribution and establish a threshold that individuals with over 50% probability are considered positive for the given region (**4**). Once thresholds are derived across all regions, we derive the spatial extent index for each participant by summing the number of positive regions across the brain. (**5**). We also apply the same methodology to the average SUVR within each aggregate composing Braak stages I and III through VI (warmer to colder colors) (**6**). To compare our spatial extent index in the cognition analyses, we also compute the average SUVR in a classic temporal meta-ROI. (**7**) CU = Cognitively unimpaired, MCI = Mild cognitive impairment, AD = Alzheimer’s disease. Figure adapted from sihnpy’s documentation (https://sihnpy.readthedocs.io/) with permission of the first author.

We also derived a more typical temporal meta-ROI^9^ and the regions composing the Braak staging scheme.^5, 6, 8^ The temporal meta-ROI was the average SUVR from key regions harboring elevated tau-PET SUVR in Alzheimer’s disease: the entorhinal cortex, the parahippocampal, inferior temporal, the middle temporal and fusiform gyri, and the amygdalae.^9^ In the Braak Staging scheme, pathology accumulation follows a predetermined order ranging from Braak I to VI until the whole cortical mantle is affected by tau (see Supplementary Table 1 for all regions included in each stage.)^6, 8^ Braak II (hippocampus) was excluded from our analyses owing to known choroid plexus off-target binding effect of the flortaucipir tau-PET tracer.^3^ We averaged the tau-PET SUVR values in bilateral regions comprising each Braak stage, following methods described previously.^5, 28^ We then applied the GMM approach, as described in Figure 1, to determine a data-driven threshold for each Braak stage. These thresholds were then applied to assign which individuals were positive on each Braak stage.

A subset of 195 participants had at least two tau-PET scans for longitudinal analyses, with 100 having three such scans. The same regional binarization of positive (score 1) or negative (score 0) using the regional cutoffs was applied to all time points.

### Neuropsychological measures

To compare the clinical implications of our regional index score vs. a typical meta-ROI analysis, we compared the association of each with composite cognitive scores for memory, executive function,^29^ language and visuo-spatial performance.^30^ The cognitive performance data were taken as the test timepoint closest in time to tau-PET. As well, we assessed cognitive decline in participants by estimating slopes of annual change for each cognitive composite score using linear mixed effects models with random slopes and intercepts. For these analyses the cognitive score at each visit was the outcome, with the exposure being time since the initial cognitive test score in ADNI. These analyses considered all ADNI visits for the whole sample, thereby maximizing the number of timepoints contributing to estimates of individual slopes.

### Statistical analyses

All statistical analyses were run using Python v3.9.2 (numpy v1.23.1; pandas v1.4.3; scipy v1.9.3; scikit-learn v1.2.1; matplotlib v3.6.3), R v4.2.0 (Packages: lme4 v1.1-30; tidyverse v1.3.1; lmerTest v3.1-3; lmtest 0.9-40; nonnest2 v0.5-5; tableone v0.13.2; patchwork v1.1.2; ggseg v1.6.5; ggnewscale v0.4.7; glue v1.6.2; MASS v7.3-59) and R Studio “Prairie Trillium” Release (1db809b8, 2022-05-16) for macOS.

### Demographics

We compared groups on their demographic information by their diagnostic status separately for Aβ+ and Aβ- participants using one-way ANOVA and Tukey post-hoc tests being used for continuous variables and chi-square tests for categorical variables.

### Cross-sectional characterization of tau

We first compared tau levels of Aβ-positive vs. Aβ-negative individuals. For the three diagnostic groups of CU, MCI, or Alzheimer’s disease dementia, we compared our spatial extent index with the temporal meta-ROI SUVR contrasting Aβ+ and Aβ- individuals within each group using Wilcoxon signed rank test. Logistic regression complemented this analysis by quantifying the probability of having a spatial extent index of at least one based on a continuous burden of Aβ pathology (centiloid values). Separately for Aβ+ and Aβ- participants, we used a heatmap to plot regions that were tau positive based on our spatial extent approach. All analyses from this point were done separately in each diagnostic group in the Aβ+ sample (tau binding being typically low in Aβ- participants). Finally, we calculated the extent to which each participant’s tau pathology was consistent with Braak staging. To do this, at each Braak stage we computed the percentage of participants who were tau-positive both at their more advanced Braak stage and at all previous stages (e.g., if a participant was positive on Braak IV, and was also positive on Braak III and I, then this participant was judged to have data in accord with Braak staging).

### Longitudinal characterization of tau

We studied change over time in the different tau-PET measures. First, we used linear mixed effect models to study the annual change of the tau measures (tau as the outcome; time since first tau scan as exposure) with random slopes and intercepts for each participant, for the temporal meta-ROI and the spatial extent index. We then used heatmaps to track the change in status between the first and last PET scan of a given participant. We used linear mixed effect models with random slopes and intercept to track the annual change in positivity and the annual change in SUVR in each brain region and plotted the regions on a template brain map. We calculated the extent to which Braak stages were followed by participants longitudinally. For each Braak stage, we computed the percentage of participants who became positive at each stage, and who were already positive or progressed in the previous Braak stages (e.g., if a participant became positive on Braak IV at their last visit and was already positive or progressed in Braak III and I, the participant followed the Braak stages).

### Tau-PET heterogeneity

We computed the overlap between the patterns of abnormal tau at baseline or over time overlapped between participants with the same diagnostic label using the Jaccard Similarity index. The index ranges from zero to one where zero indicates that not a single positive region overlaps between participants, and one indicates that all positive regions between two participants perfectly overlap. We then averaged the values so that each participant would be left with a single value representing, on average, how similar their tau positivity pattern was to the rest of their diagnostic group at the whole brain level. Analyses were always restricted to individuals with at least one positive region.

### Associations with cognition

We studied the association between our tau spatial index measures at baseline and the cognitive performance at the time of the PET, and the cognitive decline (slope) across all available cognitive visits using linear models. Beta, standardized beta, *P-*values and model fit (*R*^2^ and *AIC*), are reported. Models were adjusted for age, sex and education and were also subjected to a false discovery rate (FDR) multiple comparison correction. Difference in model fit between different tau measures were assessed using Vuong’s closeness test (i.e., non-nested likelihood ratio test).^31^

### Supplementary analyses

We assessed the association between tau uptake and cognitive performance in each of the 70 brain regions. Tau SUVR in each region was associated with cognitive performance and cognitive decline for each diagnostic group, controlling for age, sex, and education. Within each group, a False Discovery Rate (FDR) correction was applied to avoid multiple comparison issues. Beta coefficients of the surviving relationships were plotted on brain templates.

### Data availability

Data used in this study come from the Alzheimer’s disease neuroimaging initiative (ADNI). Investigators interested in obtaining the data can apply for access on ADNI’s website: https://adni.loni.usc.edu/. The code used to compute the spatial extent measures is publicly available in *sihnpy* as of version v0.2, a Python package freely available for download (https://sihnpy.readthedocs.io/). The code used for the statistical analyses and for the figures is also made available freely on Github (https://github.com/villeneuvelab/projects).

## Results

### Participants

A total of 1,370 tau scans from 832 unique participants had at least one Aβ- and tau-PET scan. At the time of the baseline tau scan, 463 participants were cognitively unimpaired (CU), 277 had mild cognitive impairment (MCI) and 92 had Alzheimer’s disease dementia. About half of the sample (51%) were female, and 34% had at least one ApoE4 allele. Participants were on average 73.56 ± 7.95 years old. Overall, 35.1% (*n* = 107) of CU individuals, 47.7% (*n* = 132) individuals with MCI, and 83.7% (*n* = 77) individuals with AD were Aβ-positive. Full demographic information is available in Table 1.

**Table 1.** – Demographic information.

|  | Aβ-negative (n=460) |  |  | Aβ-positive (n=372) |  |  |
| --- | --- | --- | --- | --- | --- | --- |
|  | CU | MCI | AD | CU | MCI | AD |
|  | (n=300) | (n=145) | (n=15) | (n=163) | (n=132) | (n=77) |
| Sex, n Females, (%) | 176 (58.67) | 56 (38.62) | 5 (33.33) | 96 (58.90) | 65 (49.24) | 32 (41.56) |
| APOE4 carriers, n (%) | 66 (22.00) | 23 (15.86) | 5 (33.33) | 73 (44.79) | 70 (53.03) | 48 (62.34) |
| Age (years) | 71.48 (7.31) | 73.72 (8.48) | 73.83 (8.43) | 74.82 (7.57) | 74.36 (7.39) | 77.35 (8.93) |
| Education (years) | 16.83 (2.30) | 16.32 (2.74) | 16.07 (2.60) | 16.64 (2.34) | 15.99 (2.49) | 15.55 (2.48) |
| Centiloid values | 4.09 (8.11) | 1.18 (10.53) | 1.63 (11.27) | 53.47 (30.83) | 75.78 (35.15) | 90.14 (32.86) |
| Memory composite score | 1.08 (0.61) <sup>b,c</sup> | 0.52 (0.62) <sup>a,c</sup> | -0.55 (0.48) <sup>a,b</sup> | 1.00 (0.62) <sup>b,c</sup> | 0.07 (0.59) <sup>a,c</sup> | -0.77 (0.57) <sup>a,b</sup> |
| Executive composite score | 1.20 (0.82) <sup>b,c</sup> | 0.61 (0.82) <sup>a,c</sup> | -0.47 (0.95) <sup>a,b</sup> | 0.92 (0.77) <sup>b,c</sup> | 0.19 (0.92) <sup>a,c</sup> | -0.79 (1.16) <sup>a,b</sup> |
| Language composite score | 0.89 (0.51) <sup>b,c</sup> | 0.52 (0.50) <sup>a,c</sup> | -0.21 (0.39) <sup>a,b</sup> | 0.75 (0.49) <sup>b,c</sup> | 0.41 (0.55) <sup>a,c</sup> | -0.18 (0.61) <sup>a,b</sup> |
| Visuospatial composite score | 0.13 (0.29) <sup>b</sup> | 0.01 (0.34) <sup>a</sup> | -0.07 (0.44) | 0.06 (0.36) <sup>c</sup> | 0.00 (0.38) <sup>c</sup> | -0.43 (0.72) <sup>a,b</sup> |
| **Longitudinal sub-sample** | **CU** | **MCI** | **AD** | **CU** | **MCI** | **AD** |
|  | **(n=96)** | **(n=40)** | **(n=10)** | **(n=90)** | **(n=66)** | **(n=39)** |
| Average number of tau PET scan per participant | 2.56 (0.87) | 2.52 (0.72) | 2.20 (0.42) | 2.68 (0.75) | 2.58 (0.66) | 2.54 (0.60) |
| Average number of cognitive visits per participant | 6.24 (3.45) | 8.75 (5.59) <sup>a, c</sup> | 3.80 (3.88) | 6.18 (3.83) | 5.55 (4.21) | 4.54 (4.02) |

a : significantly different from CU group, b: significantly different from MCI group, c: significantly different from AD group. Values correspond to mean (standard deviation) unless otherwise specified. APOE4 positivity corresponds to having at least 1 e4 allele. Statistical tests were performed within each of Aβ-negative and Aβ-positive groups.

In the Aβ-positive sample, 12.1% (*n* = 56) of CU participants, 36.1% (*n* = 100) of MCI and 73.9% (*n* = 68) of Alzheimer’s disease patients had at least one region of tau positivity (Fig. 2A, heatmap in Fig. 3A). In the Aβ-negative sample, a small percentage of participants had at least one tau-positive region (heatmap in Supplementary Fig. 1B and Supplementary Fig. 2) and had lower tau SUVR in the temporal meta-ROI (Supplementary Fig. 2). Every increase of 1 Aβ centiloid unit increased the risk of having at least one brain region with abnormal tau tracer uptake abnormal by 4% (Fig. 2B). Considering these findings, and our focus on tau pathology, we restricted the rest of the main analyses to Aβ-positive individuals (*n* = 372).

**Figure 2.**
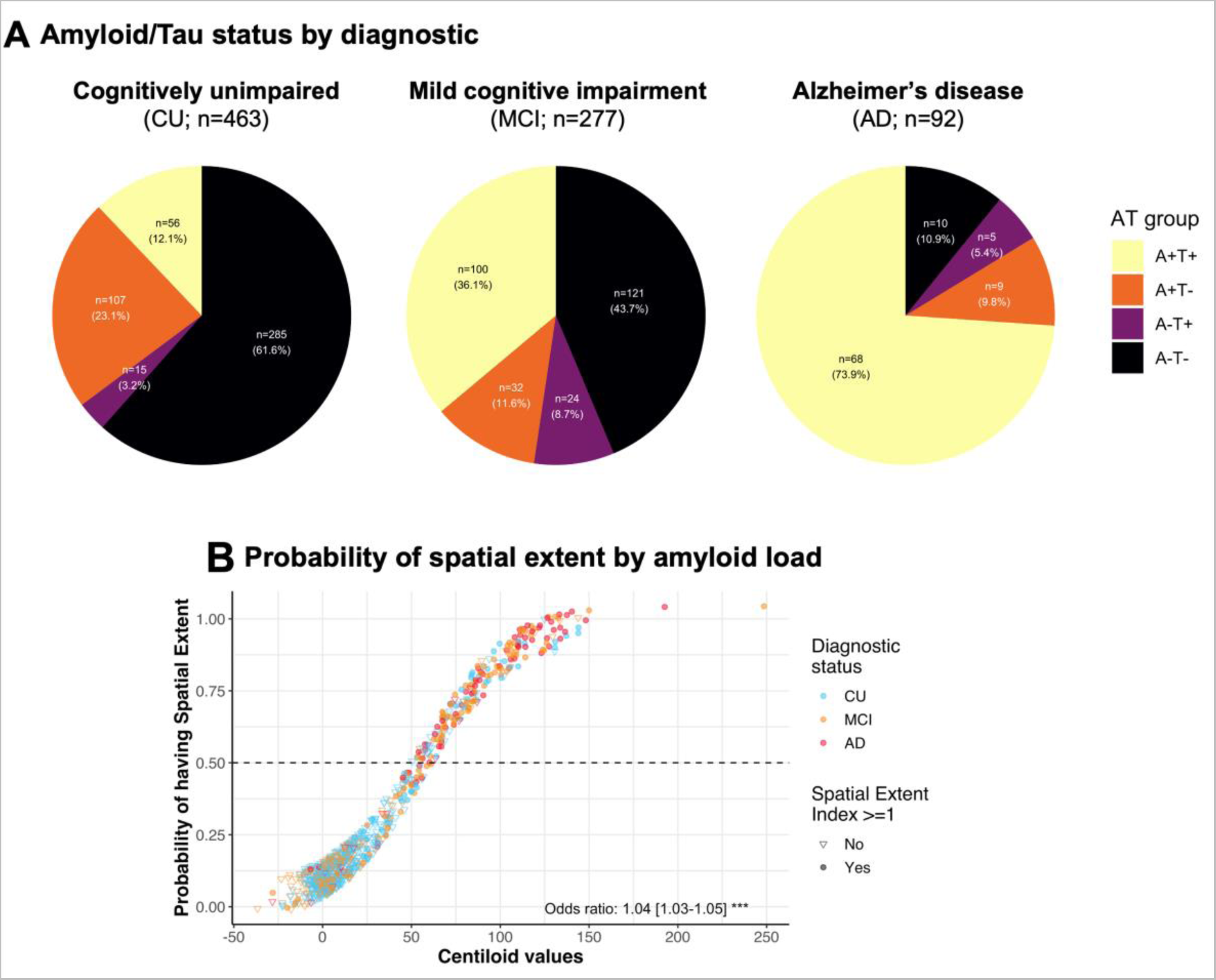
Amyloid and tau status in the cohort. (**A**) Aβ/tau (AT) status in the included participants from ADNI. Aβ positivity was established using ADNI’s tracer-specific recommendations for both florbetapir and florbetaben. Tau positivity was defined as having at least one region positive for tau pathology (spatial extent index of one and above). (**B**) Scatterplot of the probability of having at least one positive tau region (i.e., spatial extent index equal to or higher than one) as a function of the Aβ load (in centiloid). The probability was extracted from a logitistic regression. Odds ratio (and confidence interval) derived from a logistic regression is presented at the bottom of the graph. Note that the points were jittered by a factor of 0.065x0.065 for visualization purposes.

**Figure 3.**
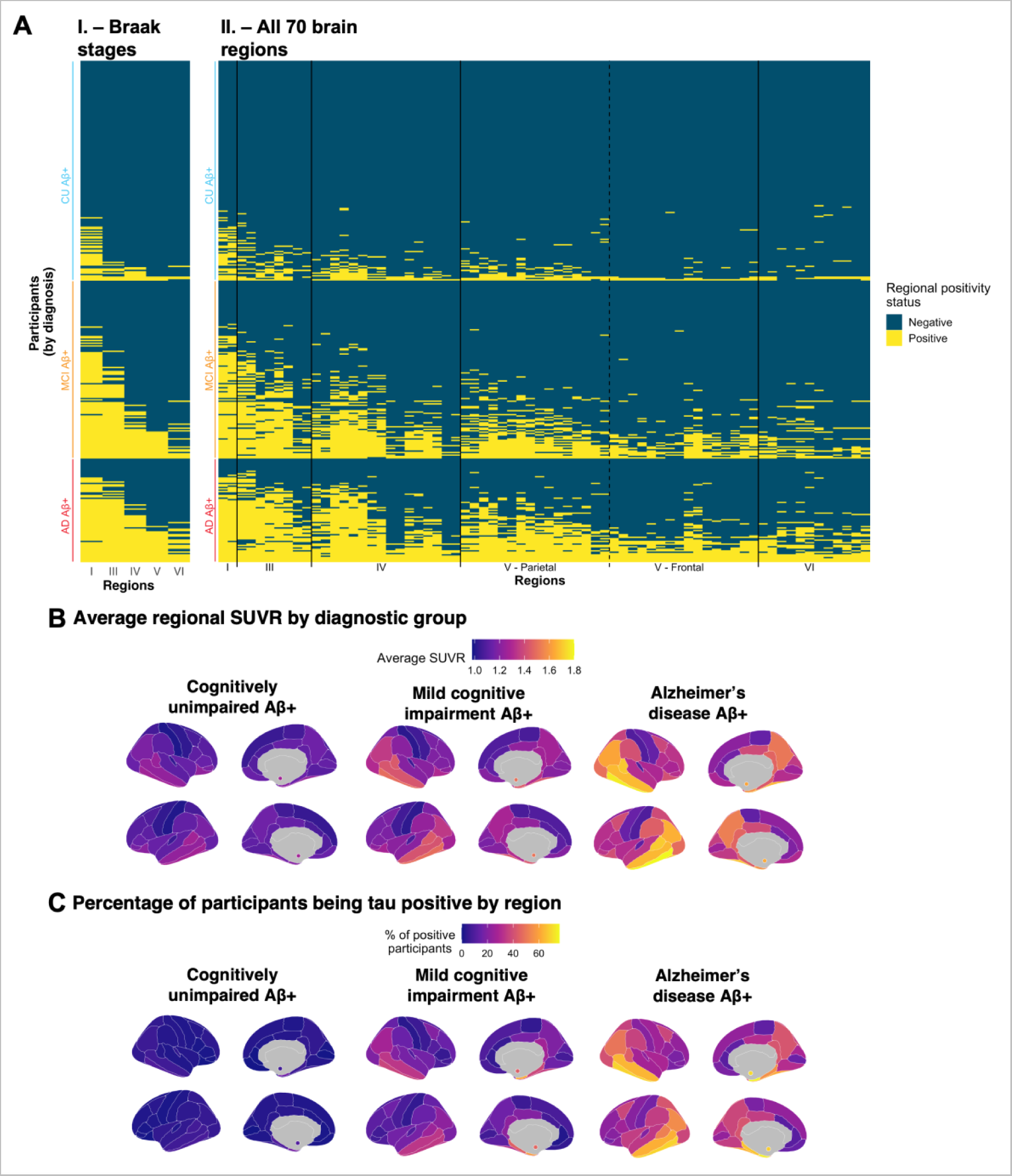
Spatial extent of abnormal tau deposition in amyloid positive participants of the ADNI cohort. (**A)** Based on the method discussed in Figure 1, abnormality thresholds were determined for each **I.** Braak stages (except stage II) and for each **II.** region of the cortical mantle and the bilateral amygdalae (70 regions). One row on the heatmap correspond to an individual participant, while each column represents a distinct cortical region. Within each diagnostic group, participants were sorted from individuals with lowest to highest spatial extent index. Regions on the x-axis in **II.** are sorted by Braak stages. (**B)** Regional average SUVR, by diagnostic status. (**C)** Brain maps representing the percentage of participants having abnormal levels of tau in each region, by diagnostic status.

### Cross-sectional tau-PET patterns

We found that, across diagnostic groups, the entorhinal cortex (Braak I) was the region most positive across Aβ-positive individuals (CU = 17.2%, MCI = 59.9%, Alzheimer’s disease = 74.7%; Fig. 3B & Supplementary Table 2). In all diagnostic groups, the five regions that were most often tau positive after the entorhinal cortex were, in order, the inferior temporal (Braak IV), the amygdalae (Braak III), the parahippocampal gyri (Braak III), the middle temporal (Braak IV) and the fusiform gyri (Braak III). All the regions above constituted the temporal meta-ROI.^9^ Similarly, we found that participants largely follow the Braak staging scheme (Fig. 3A): across all Braak stages up to and including Braak V, over 91% of participants positive on any given Braak stage were also positive on all previous Braak stages.

### Longitudinal tau-PET patterns

We repeated the analyses in our longitudinal sample (*n* = 195). Specifically, we assessed whether participants becoming positive in a Braak stage at their last tau scan were either already positive in preceding Braak stages or also progressed in previous stages during the follow-up period.

We quantified which brain regions were negative at baseline and became positive over time (progressor), were positive at baseline and became negative over time (regressor), were positive at both visits (stable positive) or were negative at both visits (stable negative). Similar to the cross-sectional results, we found that participants largely followed the Braak staging scheme (Fig. 4A): across all Braak stages up to and including Braak V, over 80% of participants who progressed on a Braak stage at follow-up were already positive or progressed on all previous Braak stages.

**Figure 4.**
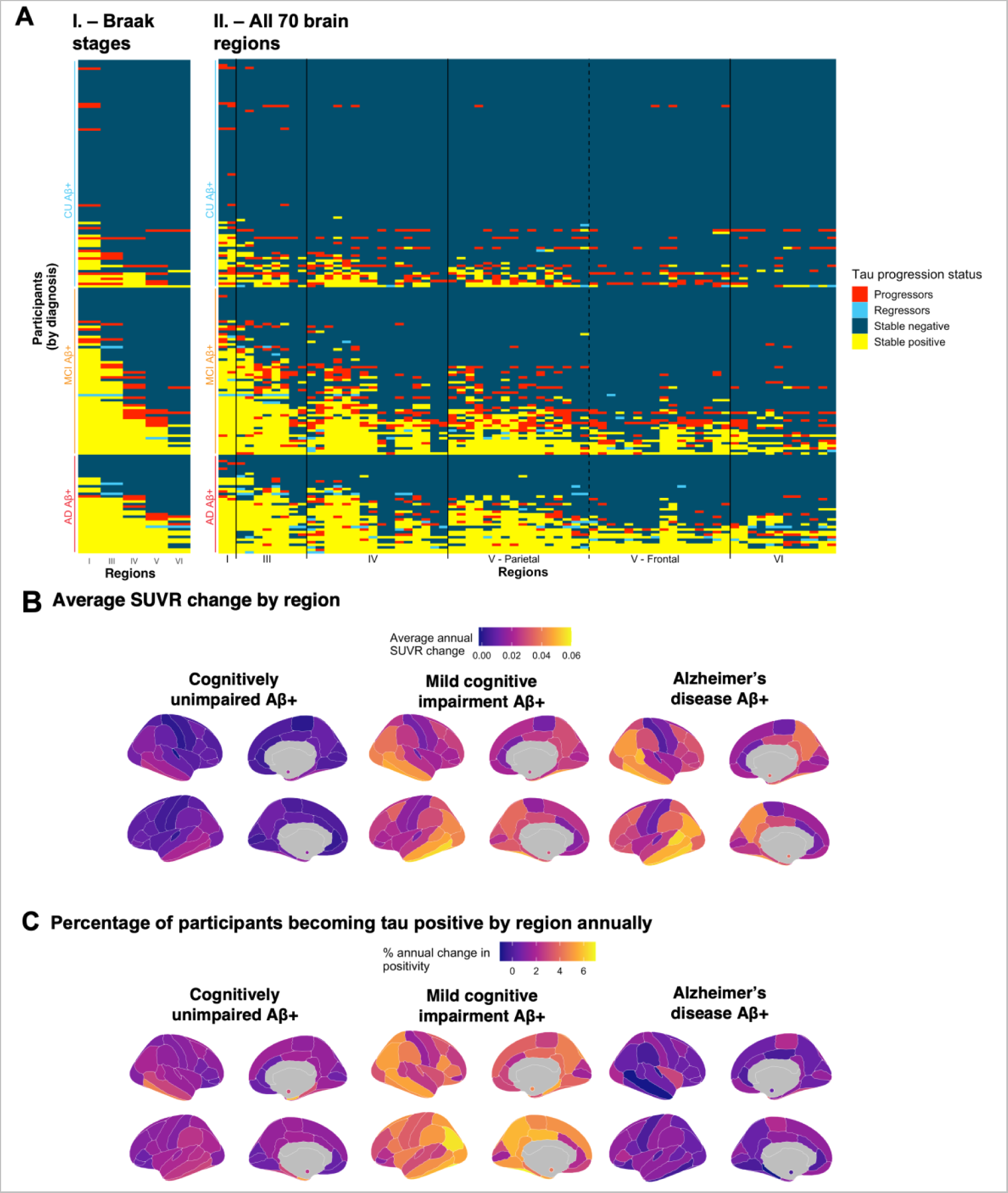
Spatial localization of abnormal tau accumulation over time in amyloid-positive participants of the ADNI cohort. (**A**) Abnormal accumulation is presented by (**I.**) Braak stages and (**II.**) all 70 individual brain regions of the Desikan atlas. Colors denote the change in the region between the baseline and the last available visit. A stable region (negative or positive; blue or yellow) did not change status during the follow-up. A progressing region (red) was originally negative and subsequently became positive over time. A regressing region (teal) was originally positive and became negative over time. (**B**) Brain maps presenting the average SUVR change per region per year. (**C**) Brain maps representing the percentage of participants becoming tau positive in each region annually. In both (**B**) and (**C**), values in the bilateral amygdalae are represented by small colored circles in the medial view of the brain, and the annual change is calculated in each region using linear mixed effect models with random slopes and intercepts. Only participants with at least three tau scans (n = 100) were kept for (**B**) and (**C**) to ensure a constant sample across the longitudinal follow-ups.

Patterns of progression across the brain however were different between clinical stages (Fig. 4; Supplementary Table 3). Specifically, CU participants mostly progressed in the entorhinal cortex (Braak I) while tau abnormality in participants with MCI progressed across the entire cortex, and few participants with Alzheimer’s disease dementia accrued additional tau abnormal regions (Fig. 4C). The annual rate of regions progressing from negative to positive was 2.5 region per year in participants with MCI, which was higher than for CU (0.968 region/year) and participants with Alzheimer’s disease dementia (0.865 region/year). (Supplementary Fig. 3B).

Few regressions from positive to negative were observed. In terms of Braak stages, 4 participants with MCI and 3 participants with Alzheimer’s disease dementia (4% of total participants) regressed from a Braak positive to a negative status (usually Braak III, V or VI). In most cases, the participants only regressed on a single Braak stage. At the regional level, thirty participants (15%) had at least one individual region regressing from positive to negative. The rate of regression was lower in CU (3%) and participants with MCI (18%) compared to participants with Alzheimer’s disease dementia (38%), which could be explained by the higher number of positive regions in these participants.

Overall, we found that participants overwhelmingly followed the Braak staging scheme, demonstrated cross-sectionally and longitudinally, excepting the very last Braak stage. However, we also show that there are substantial individual differences in abnormal regions at baseline and in the regional progression of tau pathology.

### Heterogeneity of regional tau abnormality

While abnormal tau accumulation followed Braak staging, regional tau abnormality within each Braak stage was heterogenous across individuals (Fig. 3A; Supplementary Table 2). To better understand the extent of the individual differences in tau abnormality patterns, we computed the overlap between the patterns of abnormality of different participants using the Jaccard coefficient index.

Across the whole brain and clinical groups, the average overlap was 0.55 (± 0.14; Supplementary Fig. 4A). In other words, only half of regions positive in a participant were positive in other participants. CU participants demonstrated the least heterogeneity with an average overlap of 0.74 (± 0.15), participants with MCI had an average overlap of 0.58 (± 0.14) and participants with Alzheimer’s disease dementia demonstrated the most heterogeneity with an average overlap of 0.46 (± 0.08).

We repeated the above analysis also to analyse whether patterns of tau abnormality progression differed across individuals. In this case, the Jaccard index gives an estimate of whether the same regions have the same progression status (i.e., progressor, regressor, stable positive, stable negative) across individuals in participants with at least one abnormal tau region.

Across the whole brain and clinical groups, the average overlap was 0.43 (± 0.13; Supplementary Fig. 4B). In other words, fewer than half of regions had the same progression pattern among participants. CU participants demonstrated the lowest heterogeneity with an average overlap of 0.54 (± 0.15). Participants with MCI had an average overlap of 0.44 (± 0.13), and participants with Alzheimer’s disease dementia demonstrated the greatest heterogeneity with an average overlap of 0.36 (± 0.07).

Overall, both at the cross-sectional and longitudinal levels, participants demonstrated substantial regional between-individual heterogeneity in tau abnormality and its progression. Heterogeneity was not equal between clinical labels, with CU participants usually presenting the least heterogeneity and participants with Alzheimer’s disease presenting the most.

### Associations with cognitive profiles and cognitive decline

Next, we examined whether the spatial extent index yield better associations with cognition than using the SUVR within the temporal meta-ROI as associations between tau and cognition may fall outside of the temporal lobe.

In CU participants, the spatial extent index was associated with the memory composite score (*standardized [std] β* = −0.20, *P* = 0.01, *R^2^_adj_* = 0.24) (Fig. 5) while the temporal meta-ROI was not (*std β* = −0.09, *P* > 0.10, *R^2^_adj_* = 0.21). The difference in model fit was not significant (*Vuong’s z* = −1.13, *P* = 0.13), however, suggesting that the spatial extent index provided only a marginally better model fit when compared to the more traditional temporal meta-ROI. In CU participants, neither the spatial extent index nor the temporal meta-ROI were associated with any other cognitive composite (executive, language or visuospatial) (Supplementary Fig. 5-7). In participants with MCI, both the spatial extent index and the temporal meta-ROI were nearly equally associated with the memory composite, and there were no differences in model fit (*Vuong’s z* = 0.01, *P* = 0.50). Similar findings were found for the language composite. However, the association between the executive composite and the spatial extent index was stronger than that with the temporal meta-ROI (*Vuong’s z* = −1.92, p = 0.027). There was no association between the spatial extent index or the meta-ROI and the visuospatial composite. In participants with Alzheimer’s disease, results were very similar to participants with MCI: spatial extent index and temporal meta-ROI were both equally associated with the memory and language composite, and the spatial extent index was more strongly associated with the executive composite than the temporal meta-ROI (*Vuong’s z* = −1.68, *P* = 0.045). There was no association between the spatial extent index or the temporal meta-ROI and the visuospatial composite. Looking at cognitive decline, we did not find any differences: both temporal meta-ROI and spatial extent index offered similar model fit for decline across cognitive composites.

**Figure 5.**
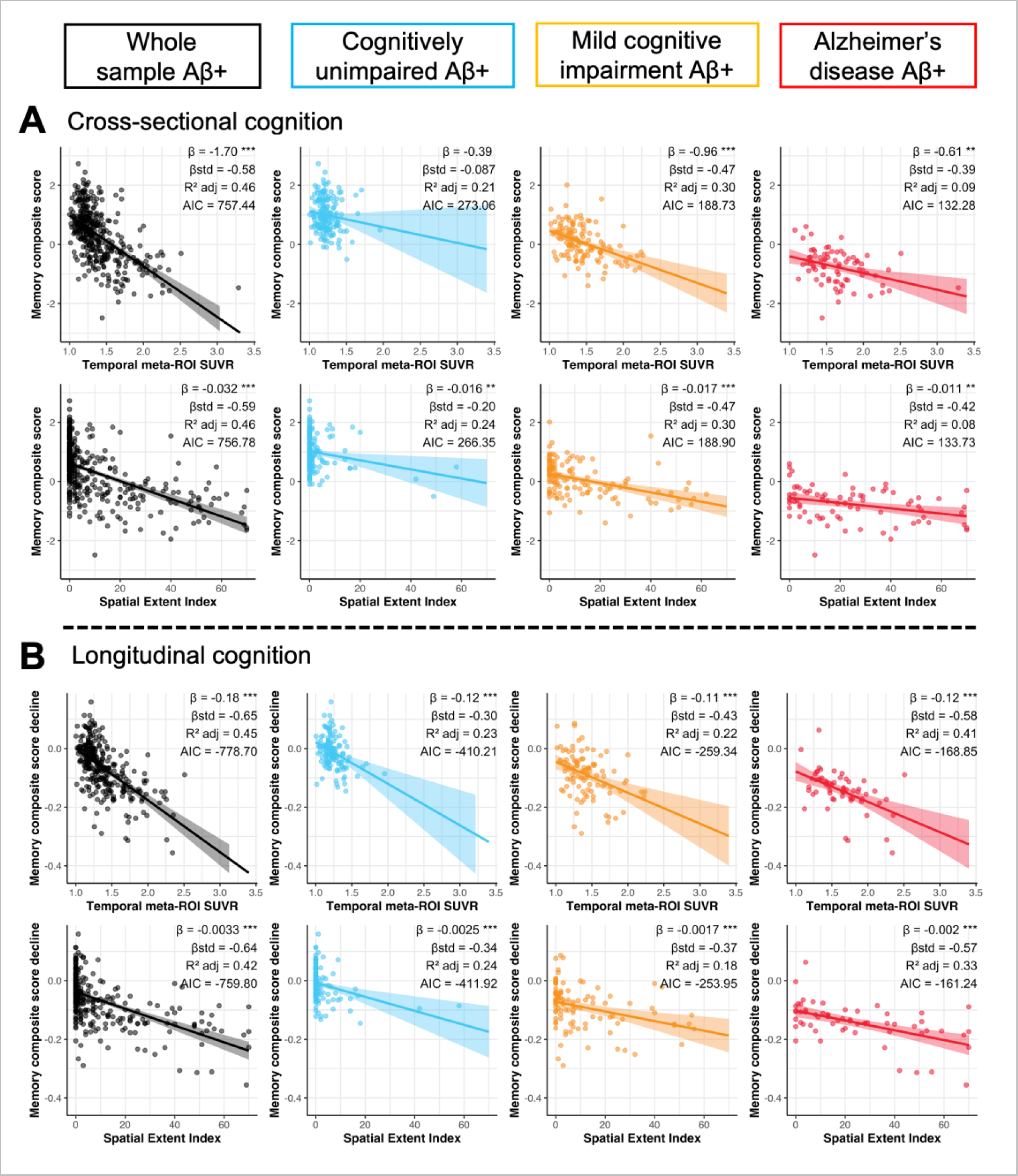
Association between tau-PET measures, and memory performance and decline. (**A**) Memory performance closest in time to the tau-PET scan and (**B**) memory decline computed across the study period were associated to both temporal meta-ROI SUVR and spatial extent index in Aβ-positive participants using linear regressions. Cognitive decline was computed for each participant with more than two cognitive timepoints using linear mixed effect models with random slopes and intercepts. In each panel, columns represent a diagnostic group (leftmost/black: whole sample, second from the left/blue: cognitively unimpaired, second from the right/orange: mild cognitive impairment, right-most/red: Alzheimer’s disease). Simple and standardized β coefficients, adjusted R^2^ and AIC, controlled for age sex and education, are shown on the graphs. P-value of models are indicated next to the simple beta coefficients. (° : P < 0.1, * : P < 0.05, ** : P < 0.01, *** P < 0.001) Results remained significant after a multiple comparison false discovery rate (FDR) correction.

In supplementary analyses, we also investigated regional associations between tau burden and cognition (Fig. 6A). In CU participants, no individual region was associated with cognitive performance on any composite score. In participants with MCI, tau levels most associated with memory were largely comprised of regions within the temporal lobe, with some weaker associations in the parietal and frontal lobes. Tau levels most associated with executive functions comprised regions across the cortex. Associations with language were almost unilaterally restricted to the left temporal lobe. No associations survived multiple correction for the visuospatial composite. Results were similar for participants with Alzheimer’s disease dementia with one exception; region-wise associations with the visuospatial composite were significant and spanned outside of the temporal lobe.

**Figure 6.**
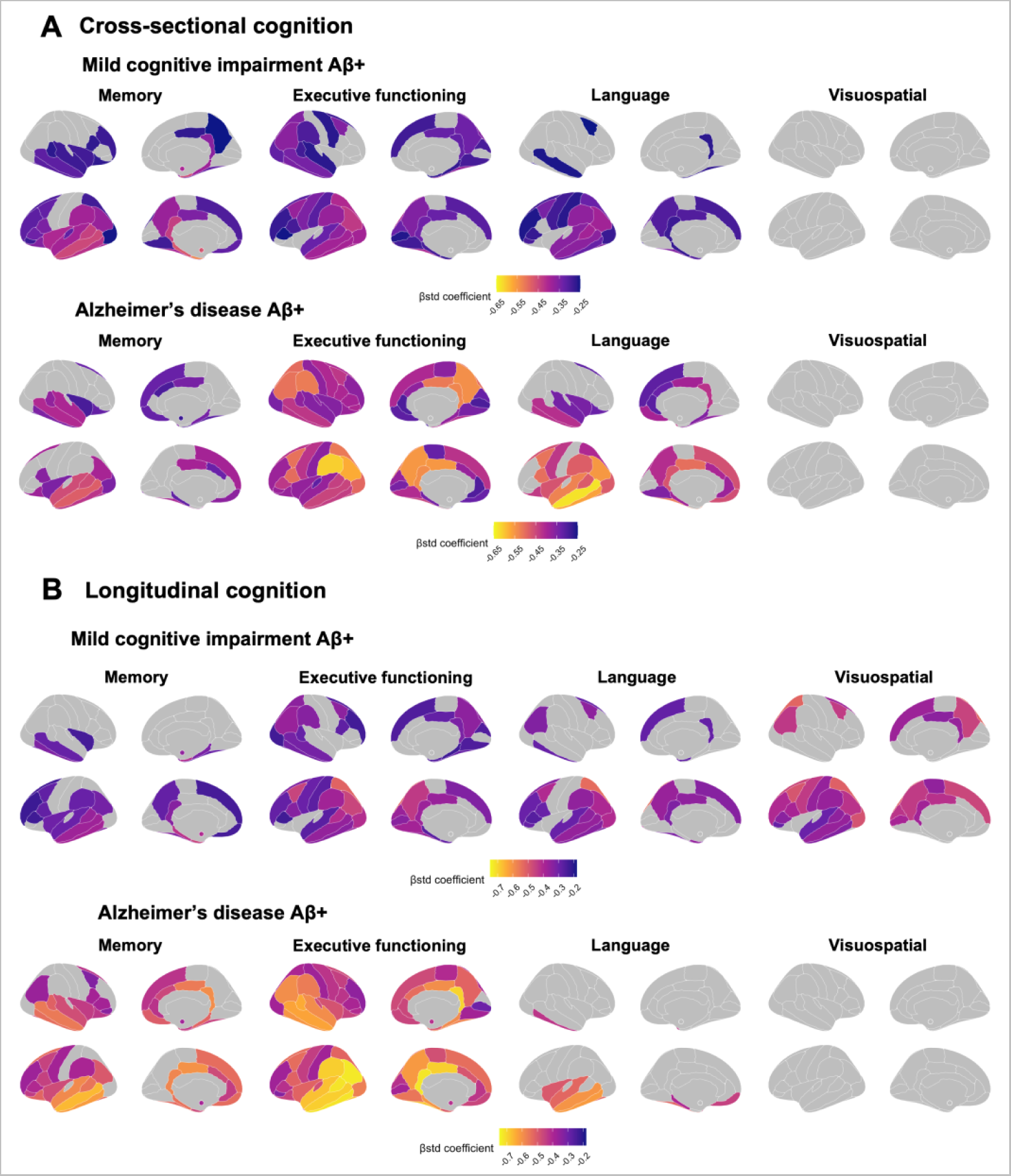
Region-wise associations between regional tau-PET SUVR and cognitive performance and decline in participants with MCI and Alzheimer’s disease. Association between tau-PET SUVR and cognitive performance (**A**) and cognitive decline (**B**) in participants with MCI and with Alzheimer’s disease across four cognitive domains (memory, executive functioning, language and visuospatial). Cognitive decline was computed for each participant with more than two cognitive timepoints using linear mixed effect models with random slopes and intercepts. The standardized β coefficients of the associations between tau-PET SUVR in a specific region and each cognition measure is displayed if it survives adjustment for age, sex and education and a multiple comparison false discovery rate (FDR) correction (P_corrected_ < 0.05).

Finally, we repeated the analyses looking at the association between baseline tau and longitudinal cognitive decline. Region-wise analyses between cognitive decline and regional tau SUVR largely replicated our findings at the cross-sectional level (Fig. 6B).

## Discussion

We found that tau spreading in late onset sporadic Alzheimer’s disease, tau pathology and follows broad stages of pathological progression (i.e., Braak stages) uniformly across individuals, with early accumulation largely constrained to temporal lobe regions. However, abnormality in cortical tau at a finer grain regional level is heterogeneous between participants, particularly as clinical symptoms progress. This effect was strongest in participants with mild cognitive impairment, who also showed the fastest region-to-region spreading of abnormal tau across the whole brain. Finally, we also found that the spatial extent index was more strongly associated with executive function performance than temporal meta-ROI SUVR in participants with MCI or Alzheimer’s disease dementia and performed on par with temporal meta-ROI SUVR in other cognitive domains. This could be due in part to the topography of the associations between executive functions and tau burden which largely spans regions outside of the temporal lobe.

In line with the literature,^6^ we found that tau pathology usually accumulates in the entorhinal cortex (Braak I) before spreading to other temporal regions (Braak III-IV)^7, 27, 32–34^ and finally large frontal and parietal regions (Braak V-VI).^8^ Similarly to previous work,^7, 27, 34, 35^ this accumulation of abnormal amounts of tau pathology was mostly restricted to participants with high levels of Aβ—as opposed to Aβ-negative participants who showed little tau abnormality. An addition from our study is that these stages are followed not just cross-sectionally, but also over time. Overall, our results recapitulate and solidify our current understand that tau pathology largely spreads following the broad Braak stages in late onset sporadic Alzheimer’s disease.

Despite these uniform broad inter-individual patterns, we found that within Braak stages, tau abnormality is regionally and inter-individually heterogenous, especially in more advanced disease stages (i.e., MCI or Alzheimer’s disease). Alzheimer’s disease is known to present many different clinical variants^36^ and heterogenous neuroimaging profiles.^15, 37^ Specifically looking at tau pathology, several “subtypes” of tau pathology have been suggested^13^ and different clinical variants of AD have also shown distinct tau deposition patterns^14, 16^. Furthermore, using individualized tau measures have been shown to better associate with future spreading of tau pathology compared to using only Braak stages, demonstrating substantial inter-individual variability.^12^ As such, it is possible that while large portion of the cortex may become abnormal following a specific sequence, regional patterns may differ between individuals. This was also suggested by a recent study which highlighted that despite tau pathology accumulating mostly in the temporal lobe, individualized regions of interest better capture change in tau overtime.^17^ Our results suggest that this heterogeneity emerges in participants with MCI. Specifically, these participants accumulated abnormal amounts of tau pathology across the entire brain faster than CU participants and participants with Alzheimer’s disease, highlighting that the heterogenous spread of pathology appears once tau has spread outside of the temporal lobe. To note, we also found that higher levels of tau pathology at baseline were associated with faster accumulation of tau pathology over time across diagnostic groups, but that the spatial extent seems to plateau at the stage of Alzheimer’s disease dementia. This suggest that there is a stage of the disease where the number of abnormal regions is reached, even though tangles (i.e., SUVR) continue to accumulate. This is somewhat contrary to Aβ pathology which seems to plateau over time at the late stage of the disease.^38^ Overall, these results suggest that fine-grain regional heterogeneity exists in tau deposition and accumulation, despite broad stages being followed uniformly, and that this heterogeneity starts to appear and accelerate in participants with MCI.

Another key finding from the study is that the extent of tau pathology across the brain is associated with cognitive performance across cognitive domains on par with tau in the temporal meta-ROI in most domains, except for executive functioning where spatial extent of tau was more strongly associated with cognition then the temporal meta-ROI. Literature in recent years has repetitively shown that tau—rather than Aβ—is the pathological hallmark most strongly associated with cognitive decline.^21^ This is also echoed by research on Alzheimer’s disease clinical variants. Previous work demonstrated that, while Aβ deposition patterns were similar across individuals from different clinical variants, tau patterns differ according to the variants, often affecting regions responsible for the main cognitive domain affected.^14, 16^ This distinct topography of tau for each cognitive domain was also found in our study: tau was associated with the memory composite mostly in the temporal and frontal lobes bilaterally, tau was associated with the executive composite across the brain and tau was associated with language mostly unilaterally to the left hemisphere. Overall, our results suggest that regional tau topography is associated with specific cognitive domains, and that leveraging the spatial extent index may uncover stronger associations between tau and executive function performance.

### Strengths and limitations

The strengths of our study include a large sample size and a large longitudinal tau-PET sample. Cognition was collected over a long follow-up period; for at least 5 years in most cases.

Our study also has some limitations to acknowledge. We staged disease progression following the clinical diagnosis as attributed by physicians from memory clinics. However, not everyone with the same clinical label may be at the same “biological” stage of the disease, i.e., two individuals with an MCI diagnosis may not have the same tau-PET patterns simply because they haven’t started to present symptoms at the same time.^13, 39^ As such, the heterogeneity observed within each clinical diagnosis could be due to participants being at more advanced disease stages. Nonetheless, our results of longitudinal tau pathology heterogeneity are reassuring: if biological staging would have been the driver of the heterogeneity in the tau patterns cross sectionally, our longitudinal results would have shown that participants had less (not more) heterogeneity.

A final major limitation is ADNI’s inclusion criteria. By design, ADNI includes participants with amnestic disease presentation.^40^ However, atypical variants of AD may not present with memory impairment at the forefront of their cognitive complaints.^36^ As such, ADNI’s sample may be by design very homogenous. This could explain why the spatial extent performed relatively similarly to the meta-ROI across cognitive composites. Despite this homogenous sample, we still found heterogenous tau patterns and diverse tau-cognition associations, and stronger association of individualized measures with executive functioning.

## Conclusion

While our study confirms that participants accumulate tau pathology following the broad Braak stages, we also demonstrate that regional accumulation is subject to significant heterogeneity— particularly as the disease progresses. This heterogeneity seems to take hold during the MCI stage, as these participants accrue more tau abnormal regions faster than both CU and participants with Alzheimer’s dementia. We also illustrate that the topography of the tau pathology is differentially associated with cognitive domains, and that using the spatial extent (i.e., tau abnormality across the brain) can lead to stronger associations with executive functioning. Taken together, our results suggest that using regional tau is important, particularly when considering participants with MCI or Alzheimer’s disease dementia, and we propose a simple method to do so.

## Data Availability

Data used in this study come from the Alzheimer's disease neuroimaging initiative (ADNI). Investigators interested in obtaining the data can apply for access on ADNI's website: https://adni.loni.usc.edu/. The code used to compute the spatial extent measures is publicly available in sihnpy as of version v0.2, a Python package freely available for download (https://sihnpy.readthedocs.io/). The code used for the statistical analyses and for the figures is also made available freely on Github (https://github.com/villeneuvelab/projects).

## Abbreviations

Aβ: Amyloid
ADNI: Alzheimer’s disease neuroimaging initiative
AIC: Akaike Information Criterion
CU: Cognitively unimpaired
FTP: Flortaucipir
GMM: Gaussian Mixture modeling
MCI: Mild cognitive impairment
PET: Positron emission tomography
ROI: Region of interest
SUVR: Standardized uptake value ratio

## Acknowledgements

The authors would like to acknowledge Valentin Ourry, Yara Yakoub, Ting Qiu, Mohammadali Javanray, Jonathan Gallego Rudolf, Bery Mohammediyan and Alfonso Fajardo for providing advice on the study research design. We would also like to acknowledge the participants of the ADNI cohort for dedicating their time and energy to collect the data used in this study and the staff working on organizing the ADNI data for making the data available to researchers worldwide.

## Funding

FSO was funded by a scholarship from the Fonds de Recherche en Santé – Québec. APB is supported by a postdoctoral fellowship from the Fonds de Recherche en Santé – Québec (298314).

Part of data collection and sharing for this project was funded by the Alzheimer’s Disease Neuroimaging Initiative (ADNI) (National Institutes of Health Grant U01 AG024904) and DOD ADNI (Department of Defense award number W81XWH-12-20012). ADNI is funded by the National Institute on Aging, the National Institute of Biomedical Imaging and Bioengineering, and through generous contributions from the following: AbbVie, Alzheimer’s Association; Alzheimer’s Drug Discovery Foundation; Araclon Biotech; BioClinica, Inc.; Biogen; BristolMyers Squibb Company; CereSpir, Inc.; Cogstate; Eisai Inc.; Elan Pharmaceuticals, Inc.; Eli Lilly and Company; EuroImmun; F. Hoffmann-LaRoche Ltdand its affiliated company Genentech, Inc.; Fujirebio; GE Healthcare; IXICO Ltd.; Janssen Alzheimer Immunotherapy Research & Development, LLC.; Johnson & Johnson Pharmaceutical Research & Development LLC.; Lumosity; Lundbeck; Merck & Co., Inc.; MesoScale Diagnostics, LLC.; NeuroRxResearch; Neurotrack Technologies; Novartis Pharmaceuticals Corporation; Pfizer Inc.; Piramal Imaging; Servier; Takeda Pharmaceutical Company; and Transition Therapeutics. The Canadian Institutes of Health Research is providing funds to support ADNI clinical sites in Canada. Private sector contributions are facilitated by the Foundation for the National Institutes of Health (www.fnih.org). The grantee organization is the Northern California Institute for Research and Education, and the study is coordinated by the Alzheimer’s Therapeutic Research Institute at the University of Southern California. ADNI data are disseminated by the Laboratory for Neuro Imaging at the University of Southern California.

## Competing interests

The authors report no competing interests.

## Supplementary material

Supplementary material is available at *Brain* online.

## Notes

### Competing Interest Statement

The authors have declared no competing interest.

### Funding Statement

FSO was funded by a scholarship from the Fonds de Recherche en Sante - Quebec. APB is supported by a postdoctoral fellowship from the Fonds de Recherche en Sante - Quebec (298314).
Part of data collection and sharing for this project was funded by the Alzheimer's Disease Neuroimaging Initiative (ADNI) (National Institutes of Health Grant U01 AG024904) and DOD ADNI (Department of Defense award number W81XWH-12-20012). ADNI is funded by the National Institute on Aging, the National Institute of Biomedical Imaging and Bioengineering, and through generous contributions from the following: AbbVie, Alzheimer's Association; Alzheimer's Drug Discovery Foundation; Araclon Biotech; BioClinica, Inc.; Biogen; BristolMyers Squibb Company; CereSpir, Inc.; Cogstate; Eisai Inc.; Elan Pharmaceuticals, Inc.; Eli Lilly and Company; EuroImmun; F. Hoffmann-LaRoche Ltdand its affiliated company Genentech, Inc.; Fujirebio; GE Healthcare; IXICO Ltd.; Janssen Alzheimer Immunotherapy Research & Development, LLC.; Johnson & Johnson Pharmaceutical Research & Development LLC.; Lumosity; Lundbeck; Merck & Co., Inc.; MesoScale Diagnostics, LLC.; NeuroRxResearch; Neurotrack Technologies; Novartis Pharmaceuticals Corporation; Pfizer Inc.; Piramal Imaging; Servier; Takeda Pharmaceutical Company; and Transition Therapeutics. The Canadian Institutes of Health Research is providing funds to support ADNI clinical sites in Canada. Private sector contributions are facilitated by the Foundation for the National Institutes of Health (www.fnih.org). The grantee organization is the Northern California Institute for Research and Education, and the study is coordinated by the Alzheimer's Therapeutic Research Institute at the University of Southern California. ADNI data are disseminated by the Laboratory for Neuro Imaging at the University of Southern California.

### Author Declarations

We used data from the Alzheimer's Disease Neuroimaging Initiative (ADNI) in this project. Approval to use ADNI data was obtained by filling an online application to the ADNI. ADNI obtained ethics approvals from all sites collecting data. The following are the ethics committees and IRB boards that provided approval: Albany Medical Center Committee on Research Involving Human Subjects Institutional Review Board, Boston University Medical Campus and Boston Medical Center Institutional Review Board, Butler Hospital Institutional Review Board, Cleveland Clinic Institutional Review Board, Columbia University Medical Center Institutional Review Board, Duke University Health System Institutional Review Board, Emory Institutional Review Board, Georgetown University Institutional Review Board, Health Sciences Institutional Review Board, Houston Methodist Institutional Review Board, Howard University Office of Regulatory Research Compliance, Icahn School of Medicine at Mount Sinai Program for the Protection of Human Subjects, Indiana University Institutional Review Board, Institutional Review Board of Baylor College of Medicine, Jewish General Hospital Research Ethics Board, Johns Hopkins Medicine Institutional Review Board, Lifespan - Rhode Island Hospital Institutional Review Board, Mayo Clinic Institutional Review Board, Mount Sinai Medical Center Institutional Review Board, Nathan Kline Institute for Psychiatric Research & Rockland Psychiatric Center Institutional Review Board, New York University Langone Medical Center School of Medicine Institutional Review Board, Northwestern University Institutional Review Board, Oregon Health and Science University Institutional Review Board, Partners Human Research Committee Research Ethics, Board Sunnybrook Health Sciences Centre, Roper St. Francis Healthcare Institutional Review Board, Rush University Medical Center Institutional Review Board, St. Joseph's Phoenix Institutional Review Board, Stanford Institutional Review Board, The Ohio State University Institutional Review Board, University Hospitals Cleveland Medical Center Institutional Review Board, University of Alabama Office of the IRB, University of British Columbia Research Ethics Board, University of California Davis Institutional Review Board Administration, University of California Los Angeles Office of the Human Research Protection Program, University of California San Diego Human Research Protections Program, University of California San Francisco Human Research Protection Program, University of Iowa Institutional Review Board, University of Kansas Medical Center Human Subjects Committee, University of Kentucky Medical Institutional Review Board, University of Michigan Medical School Institutional Review Board, University of Pennsylvania Institutional Review Board, University of Pittsburgh Institutional Review Board, University of Rochester Research Subjects Review Board, University of South Florida Institutional Review Board, University of Southern, California Institutional Review Board, UT Southwestern Institution Review Board, VA Long Beach Healthcare System Institutional Review Board, Vanderbilt University Medical Center Institutional Review Board, Wake Forest School of Medicine Institutional Review Board, Washington University School of Medicine Institutional Review Board, Western Institutional Review Board, Western University Health Sciences Research Ethics Board, and Yale University Institutional Review Board.

