## Supplementary Table 1 for "Tau accumulation and its spatial progression across the Alzheimer’s disease spectrum"

### *Supplementary material*

Frédéric St-Onge[1,2], Marianne Chapleau[3], John CS Breitner[2,4], Sylvia Villeneuve[2,4,5†], and Alexa Pichet Binette[6,†], for the Alzheimer's Disease Neuroimaging Initiative*

[†]**These authors contributed equally to this work.**

A**uthor affiliations:**

1 Integrated Program in Neuroscience, Faculty of medicine, McGill University, Montreal, Qc, H3A 2B4, Canada

2 Research Center of the Douglas Mental Health University Institute, Montreal, Qc, H4H 1R3, Canada

3 Faculty of medicine, University of California San Francisco, San Francisco, CA, 94143, United-States

4 Department of psychiatry, Faculty of medicine, McGill University, Montreal, QC, H3A 1Y2, Canada

5 McConnell Brain Imaging Centre, Montreal Neurological Institute, Montreal, QC, H3A 2B4, Canada

6 Clinical Memory Research Unit, Faculty of Medicine, Lund University, Malmö, 205 02, Sweden

*Data used in preparation of this article were obtained from the Alzheimer's Disease Neuroimaging Initiative (ADNI) database (adni.loni.usc.edu). As such, the investigators within the ADNI contributed to the design and implementation of ADNI and/or provided data but did not

participate in analysis or writing of this report. A complete listing of ADNI investigators can be found at:

http://adni.loni.usc.edu/wp-content/uploads/how_to_apply/ADNI_Acknowledgement_List.pdf

Correspondence to: Dr. Alexa Pichet Binette

Clinical Memory Research Unit

Biomedical Center, Lund University,

Sölvegatan 19, 223 62 Lund, Sweden

&

Dr. Sylvia Villeneuve

Douglas Mental Health University Institute

Perry Pavilion

6875 Boulevard LaSalle, Montreal, QC, H4H 1R3, Canada

**Supplementary Figure 1**

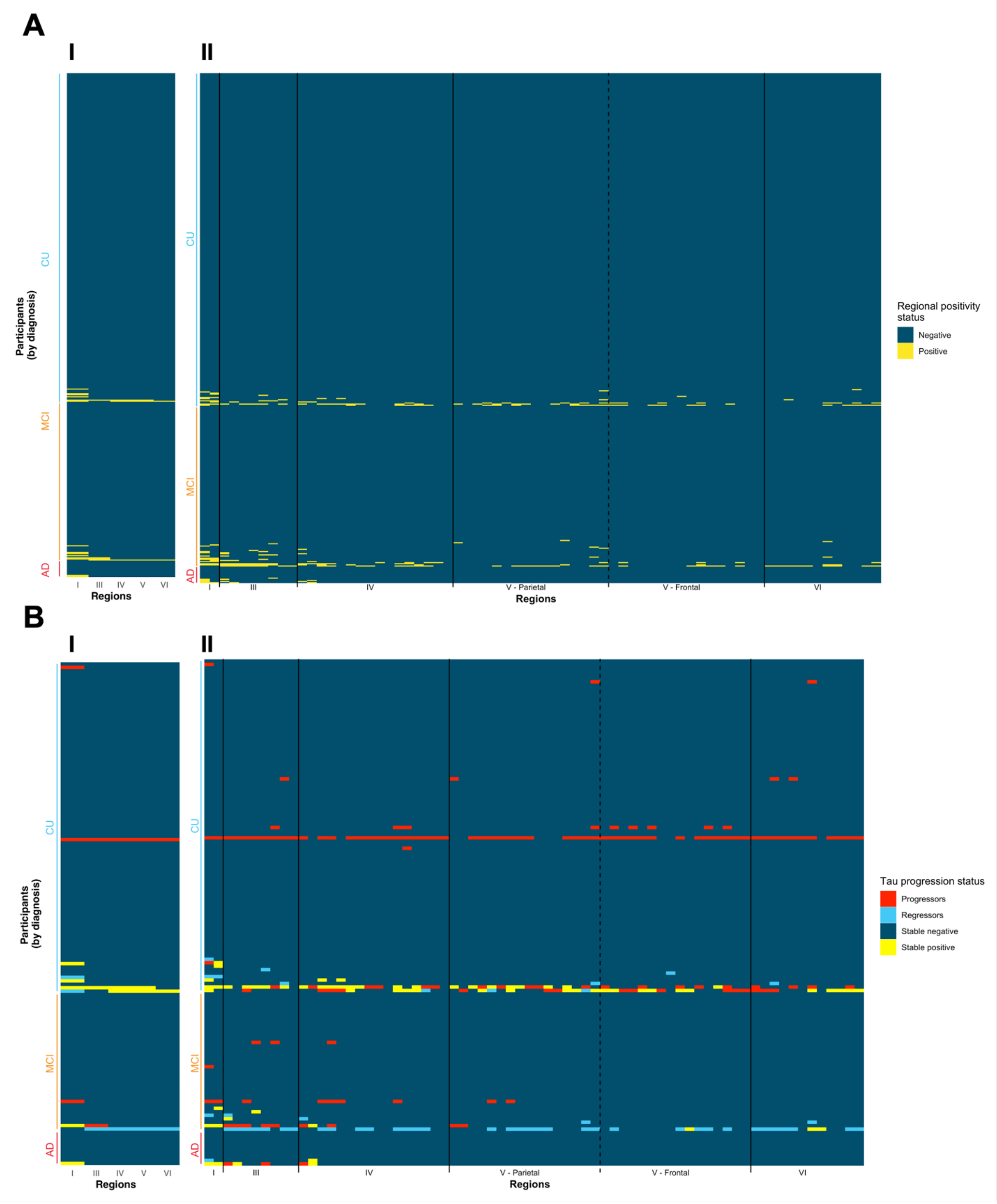


**Supplementary Figure 1 Spatial extent of abnormal tau deposition and accumulation in amyloid negative participants of the ADNI cohort.** (**A**) Based on the method discussed in Figure

1, abnormality thresholds were determined for each (**I.**) Braak stages (except stage II) and for each (**II.**) region of the cortical mantle and the bilateral amygdalae (70 regions). One row on the heatmap correspond to an individual participant, while each column represents a distinct cortical region. Within each diagnostic group, participants were sorted from individuals with lowest to highest spatial extent index. Regions on the x-axis in **II.** are sorted by Braak stages. (**B**) Abnormal accumulation is presented by (**I.**) Braak stages and (**II.**) all 70 individual brain regions of the Desikan atlas. Colors denote the change in the region between the baseline and the last available visit. A stable region (negative or positive; blue or yellow) did not change status during the follow-up. A progressing region (red) was originally negative and subsequently became positive over time. A regressing region (teal) was originally positive and became negative over time.

**Supplementary Figure 2**

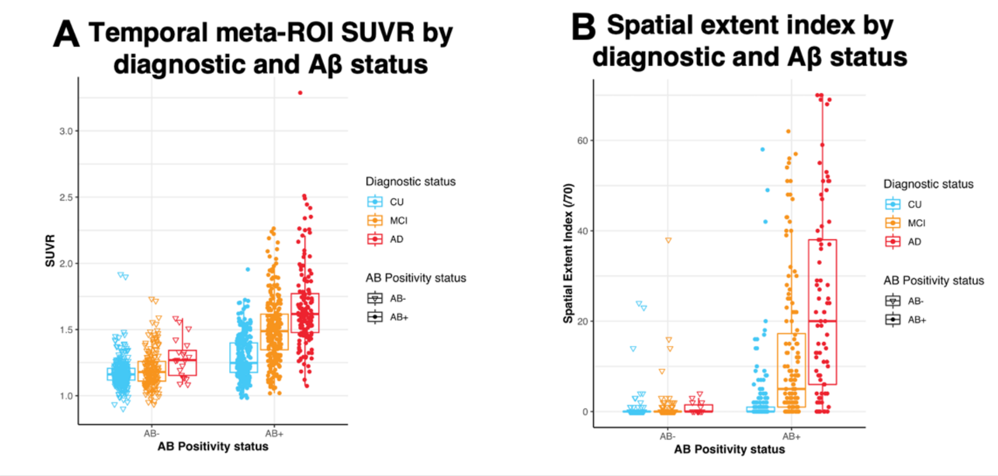


**Supplementary Figure 2 Tau measures by amyloid and clinical status. (A)** Average tau SUVR within the temporal meta-ROI by amyloid positivity and diagnostic status. (**B**) Spatial extent index (i.e., number of tau abnormal regions) by amyloid positivity and diagnostic status. In both panels, ANOVA were used to compare tau measures between Aβ- and Aβ+ participants (e.g., Aβ- compared to Aβ+ cognitively unimpaired participants). As all analyses yielded that Aβ+ had more tau—across all diagnostic groups—at $p < 0.001$ significance, we did not plot the model significance on the figure.

**Supplementary Figure 3**

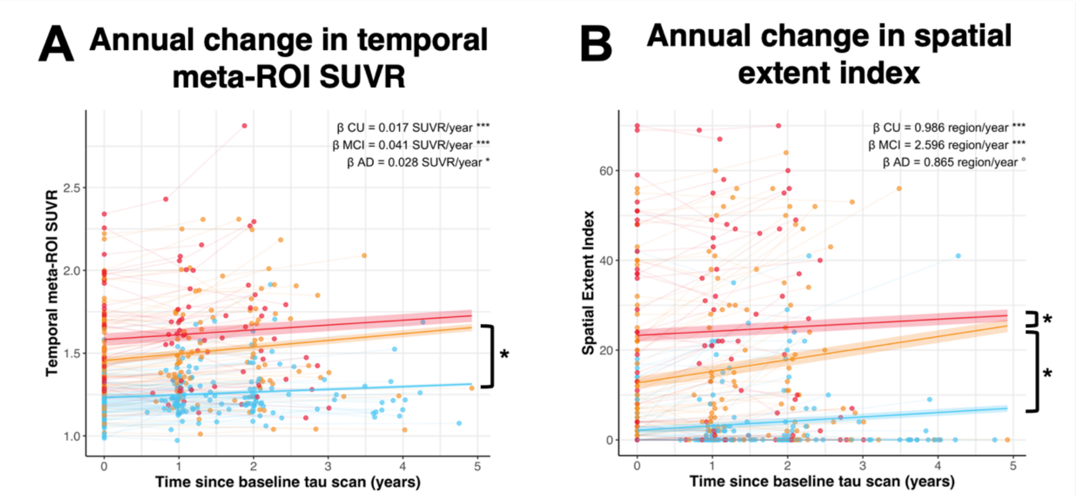


**Supplementary Figure 3 Annual change in tau-PET measures.** Annual change of (**A**) standardized uptake value ratio (SUVR) in the temporal meta-ROI and annual change of (**B**) spatial extent index in CU (blue), MCI (orange) and AD (red). Rates of annual change, computed with linear mixed models, are presented at the top right corner of the graphs. Models' significance when controlling for age, sex and education are denoted by stars next to the rate. Brackets and stars between two slopes denote a significant group difference in the rate of change (* = $P < 0.05$).

**Supplementary Figure 4**

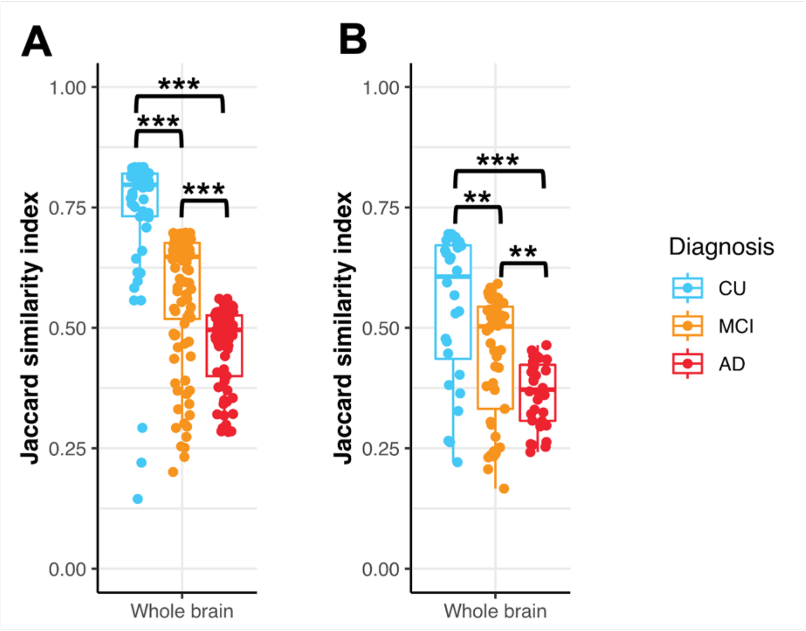


**Supplementary Figure 4 Heterogeneity in tau-PET spatial extent at baseline and longitudinally.** (**A**) Across the whole brain, we computed how, on average, the patterns of tau abnormality (i.e., positivity for specific sets of brain regions) overlapped between participants of the same diagnostic group using the Jaccard similarity index. An index closer to one means a bigger overlap on average between participants in terms of regions that are positive, while an index closer to zero means more heterogeneity on average between participants. For each diagnostic group, we only retained participants who had at least one tau positive region. (**B**). Across the brain we computed how, on average, the patterns of change in tau abnormality (i.e., stability, progression, or regression for specific sets of brain regions) overlapped between participants of the same diagnostic group using the same method described in (**A**)**.** In both (**A**) and (**B**), difference in average similarity was compared using Kruskal-Wallis tests. Post-hoc Dunn tests (with Bonferroni correction) were conducted when the result was significant. * = $P < 0.05$, ** = $P < 0.01$, *** = $P < 0.001$. CU = Cognitively unimpaired, MCI = Mild cognitive impairment, AD = Alzheimer's disease

**Supplementary Figure 5**

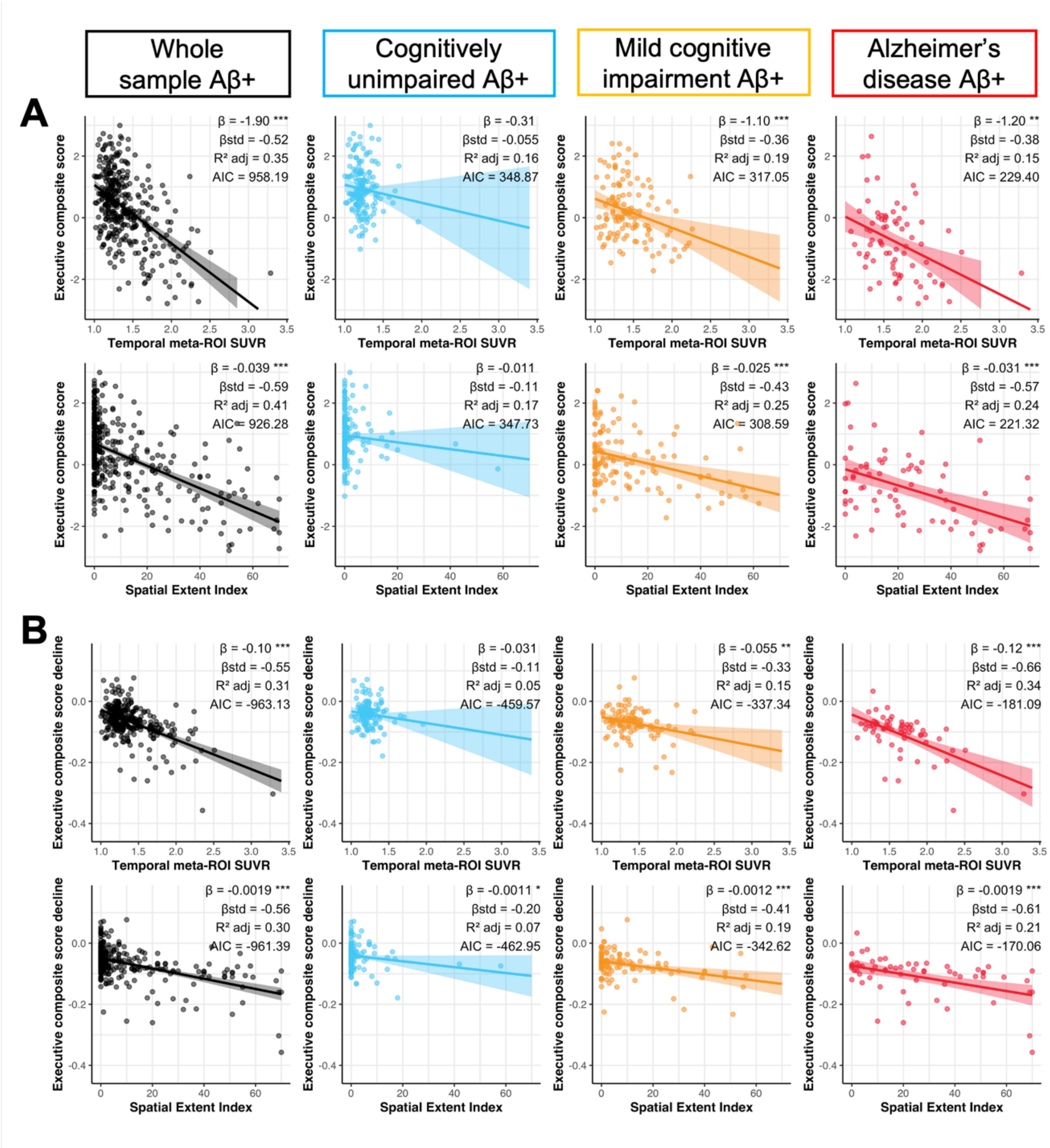


**Supplementary Figure 5 Association between tau-PET measures, and executive functioning performance and decline.** (**A**) Executive functioning performance closest in time to the tau-PET scan and (**B**) executive functioning decline computed across the study period were associated to both temporal meta-ROI SUVR and spatial extent index in Aβ+ participants using linear regressions. Cognitive decline was computed for each participant with more than two cognitive timepoints using linear mixed effect models with random slopes and intercepts. In each panel, columns represent a diagnostic group (leftmost/black: whole sample, second from the left/blue: cognitively unimpaired, second from the right/orange: mild cognitive impairment, right-most/red: Alzheimer's disease). Simple and standardized $\beta$ coefficients, *adjusted* $R^2$ and *AIC*, controlled for age sex and education, are shown on the graphs. *P*-value of models are indicated next to the simple beta coefficients. (° : $P < 0.1$, * : $P < 0.05$, ** : $P < 0.01$, *** $P < 0.001$) Results remained significant after a multiple comparison false discovery rate (FDR) correction.

**Supplementary Figure 6**

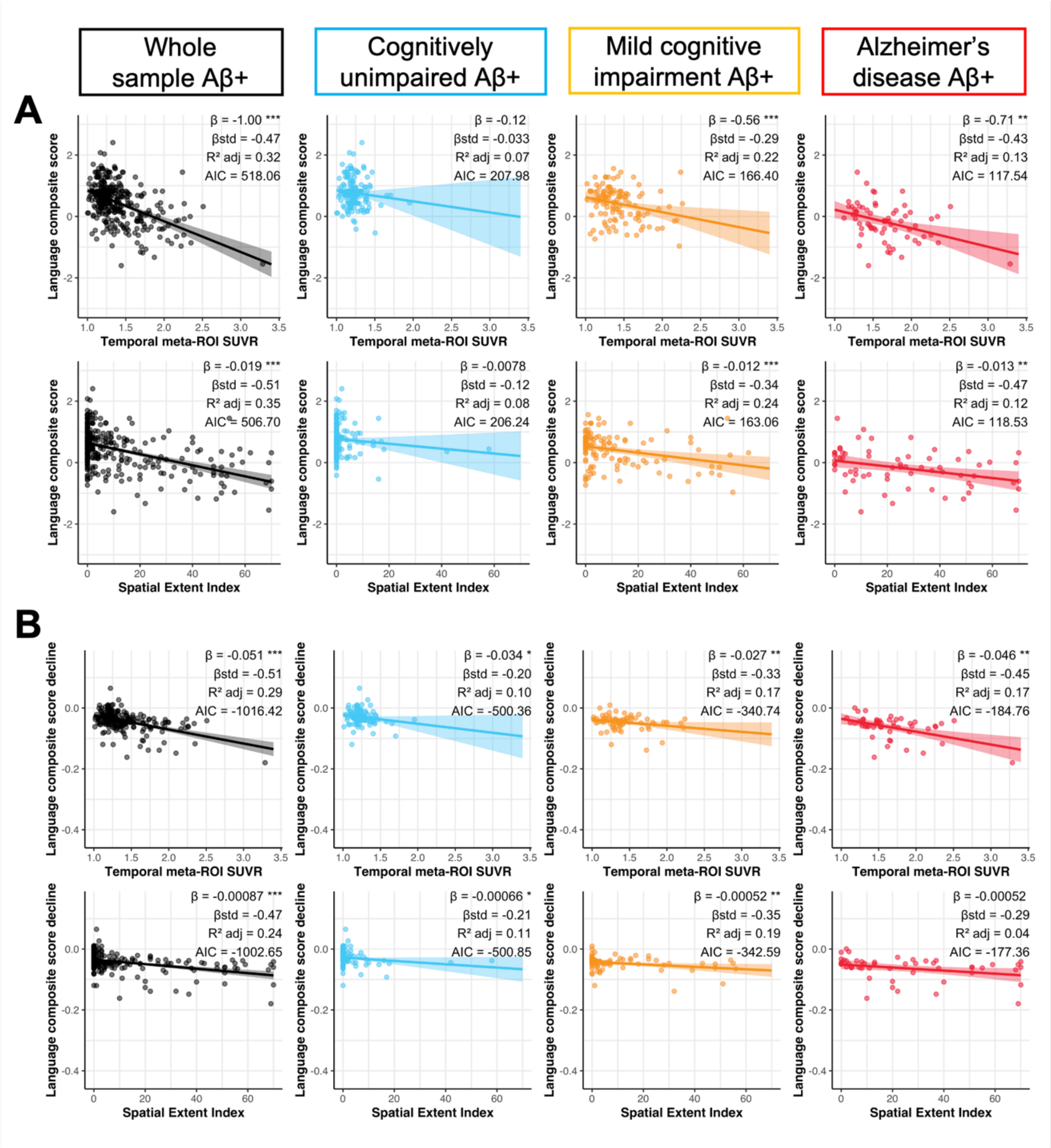


**Supplementary Figure 6 Association between tau-PET measures, and language performance and decline.** (**A**) Language performance closest in time to the tau-PET scan and (**B**) language decline computed across the study period were associated to both temporal meta-ROI SUVR and spatial extent index in Aβ+ participants using linear regressions. Cognitive decline was computed for each participant with more than two cognitive timepoints using linear mixed effect models with random slopes and intercepts. In each panel, columns represent a diagnostic group (leftmost/black: whole sample, second from the left/blue: cognitively unimpaired, second from the right/orange: mild cognitive impairment, right-most/red: Alzheimer's disease). Simple and standardized *β* coefficients, *adjusted* $R^2$ and *AIC*, controlled for age sex and education, are shown on the graphs.

$P$-value of models are indicated next to the simple beta coefficients. (° : $P < 0.1$, * : $P < 0.05$, ** : $P < 0.01$, *** $P < 0.001$) Results remained significant after a multiple comparison false discovery rate (FDR) correction.

**Supplementary Figure 7**

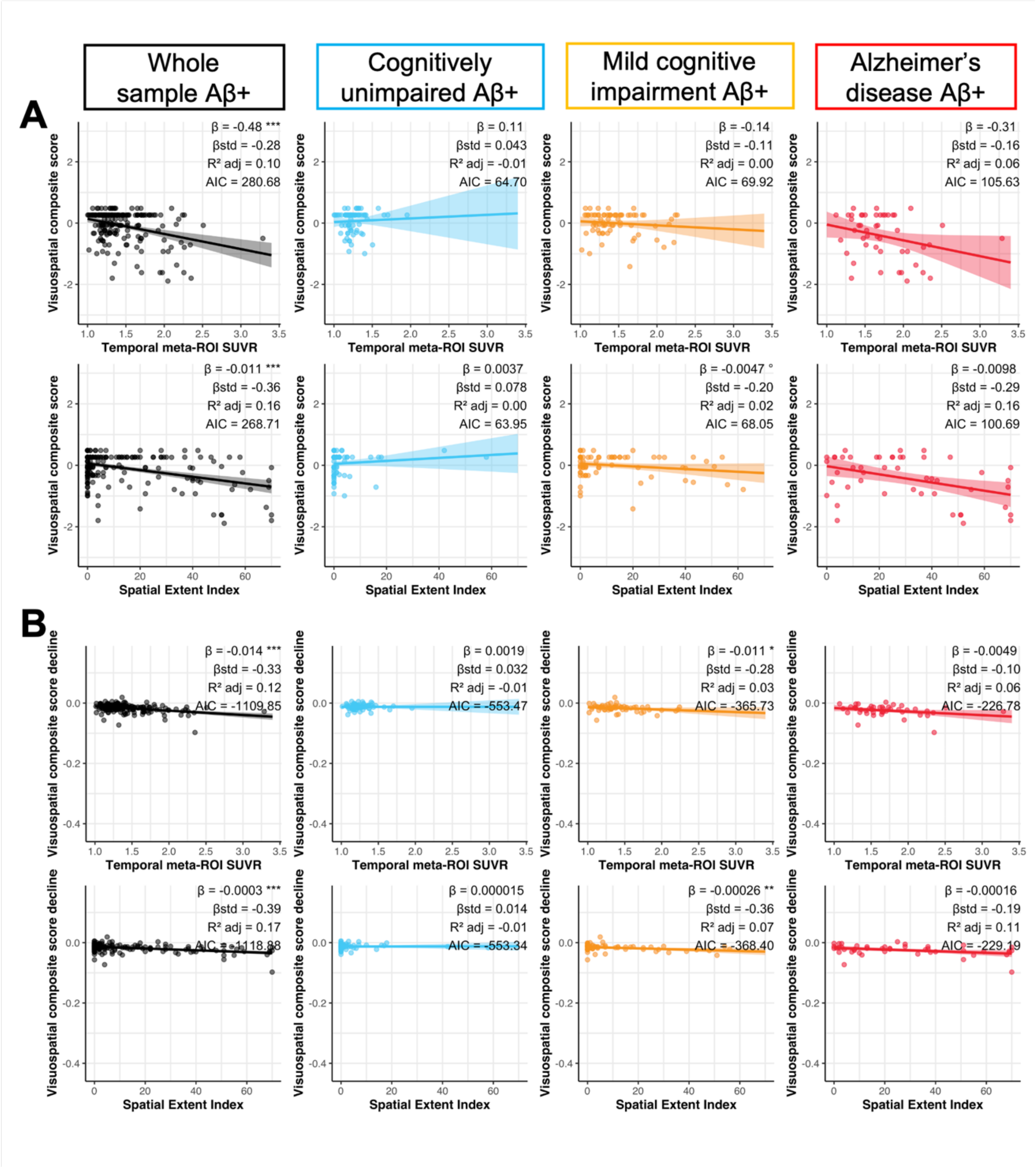

**Supplementary Figure 7 Association between tau-PET measures, and visuospatial performance and decline.** (**A**) Visuospatial performance closest in time to the tau-PET scan and (**B**) visuospatial decline computed across the study period were associated to both temporal meta-ROI SUVR and spatial extent index in Aβ+ participants using linear regressions. Cognitive decline was computed for each participant with more than two cognitive timepoints using linear mixed effect models with random slopes and intercepts. In each panel, columns represent a diagnostic group (leftmost/black: whole sample, second from the left/blue: cognitively unimpaired, second from the right/orange: mild cognitive impairment, right-most/red: Alzheimer's disease). Simple and standardized $\beta$ coefficients, *adjusted* $R^2$ and *AIC*, controlled for age sex and education, are shown on the graphs. *P*-value of models are indicated next to the simple beta coefficients. (° : $P < 0.1$, * : $P < 0.05$, ** : $P < 0.01$, *** $P < 0.001$) Results remained significant after a multiple comparison false discovery rate (FDR) correction.

**Supplementary Table 1 – Regional thresholds of tau positivity**

| Braak stage | Region | Threshold value (SUVR) LH | RH |
|---|---|---|---|
| I | entorhinal | 1.344 | 1.349 |
| III | amygdala | 1.498 | 1.501 |
| | fusiform | 1.434 | 1.421 |
| | parahippocampal | 1.319 | 1.330 |
| | lingual | 1.328 | 1.322 |
| IV | temporalpole | 1.395 | 1.402 |
| | inferiortemporal | 1.464 | 1.440 |
| | middletemporal | 1.423 | 1.421 |
| | isthmuscingulate | 1.360 | 1.360 |
| | caudalanteriorcingulate | 1.342 | 1.372 |
| | insula | 1.332 | 1.346 |
| | posteriorcingulate | 1.362 | 1.384 |
| | rostralanteriorcingulate | 1.414 | 1.391 |
| $V_1$ | lateraloccipital | 1.390 | 1.390 |
| | inferiorparietal | 1.420 | 1.403 |
| | superiortemporal | 1.333 | 1.347 |
| | bankssts | 1.490 | 1.525 |
| | precuneus | 1.407 | 1.387 |
| | superiorparietal | 1.317 | 1.310 |
| | supramarginal | 1.361 | 1.349 |
| | transversetemporal | 1.260 | 1.209 |
| $V_2$ | parsopercularis | 1.362 | 1.381 |
| | parsorbitalis | 1.486 | 1.483 |
| | parstriangularis | 1.424 | 1.423 |
| | frontalpole | 1.328 | 1.357 |
| | caudalmiddlefrontal | 1.320 | 1.292 |
| | lateralorbitofrontal | 1.491 | 1.534 |
| | medialorbitofrontal | 1.462 | 1.425 |
| | rostralmiddlefrontal | 1.349 | 1.342 |
| | superiorfrontal | 1.280 | 1.266 |
| VI | pericalcarine | 1.382 | 1.400 |
| | cuneus | 1.353 | 1.369 |
| | paracentral | 1.258 | 1.292 |
| | postcentral | 1.237 | 1.229 |
| | precentral | 1.244 | 1.225 |

LH = Left hemisphere, RH = Right hemisphere, SUVR = Standardized uptake value ratio. $V_2$ represents Braak V region in the frontal lobe while $V_1$ regroups the remaining Braak V regions.

**Supplementary Table 2 – Regional tau abnormality across regions of interest**

| | | **CU Aβ+** (n = 163) | | **MCI Aβ+** (n = 132) | | **AD Aβ+** (n = 77) | | **CU Aβ-** (n = 300) | | **MCI Aβ-** (n = 145) | | **AD Aβ-** (n = 15) | |
|---|---|---|---|---|---|---|---|---|---|---|---|---|---|
| **Braak stage** | **Region** | **LH** | **RH** | **LH** | **RH** | **LH** | **RH** | **LH** | **RH** | **LH** | **RH** | **LH** | **RH** |
| I | entorhinal * | 17.8 | 16.6 | 58.3 | 61.4 | 74 | 75.3 | 2 | 2 | 6.2 | 4.8 | 26.7 | 6.7 |
| III | amygdala * | 9.8 | 6.7 | 46.2 | 43.9 | 67.5 | 70.1 | 0.3 | 0.3 | 4.8 | 2.8 | 13.3 | 6.7 |
| | fusiform * | 5.5 | 5.5 | 31.8 | 35.6 | 62.3 | 61 | 0.3 | 0.3 | 2.1 | 2.8 | 0 | 0 |
| | parahippocampal * | 9.8 | 6.1 | 44.7 | 37.9 | 66.2 | 55.8 | 1 | 0.3 | 4.1 | 2.8 | 0 | 6.7 |
| | lingual | 1.8 | 2.5 | 12.1 | 15.9 | 35.1 | 31.2 | 0.7 | 0 | 0.7 | 0.7 | 0 | 0 |
| IV | temporalpole | 2.5 | 3.7 | 23.5 | 18.9 | 48.1 | 41.6 | 0.7 | 0.3 | 2.8 | 1.4 | 6.7 | 13.3 |
| | inferiortemporal * | 8.6 | 9.8 | 34.1 | 37.1 | 68.8 | 68.8 | 0.7 | 0.3 | 2.1 | 1.4 | 0 | 0 |
| | middletemporal * | 6.7 | 6.7 | 33.3 | 33.3 | 62.3 | 63.6 | 0.7 | 0.7 | 0 | 0.7 | 0 | 0 |
| | isthmuscingulate | 3.1 | 4.3 | 22 | 26.5 | 41.6 | 41.6 | 0.3 | 0 | 0.7 | 0.7 | 0 | 0 |
| | caudalanteriorcingulate | 1.8 | 1.8 | 7.6 | 5.3 | 11.7 | 13 | 0 | 0 | 0.7 | 0 | 0 | 0 |
| | insula | 4.3 | 2.5 | 16.7 | 16.7 | 35.1 | 28.6 | 0.7 | 0.7 | 1.4 | 2.1 | 0 | 0 |
| | posteriorcingulate | 3.1 | 2.5 | 19.7 | 17.4 | 35.1 | 31.2 | 0.7 | 0.3 | 1.4 | 1.4 | 0 | 0 |
| | rostralanteriorcingulate | 1.2 | 0.6 | 2.3 | 4.5 | 11.7 | 11.7 | 0 | 0 | 0.7 | 0 | 0 | 0 |
| $V_1$ | lateraloccipital | 4.9 | 4.3 | 18.2 | 18.9 | 37.7 | 40.3 | 0.3 | 0 | 0.7 | 0 | 0 | 0 |
| | inferiorparietal | 4.9 | 6.1 | 22 | 27.3 | 54.5 | 49.4 | 0.3 | 0.7 | 0 | 0 | 0 | 0 |
| | superiortemporal | 4.3 | 1.8 | 17.4 | 17.4 | 33.8 | 27.3 | 0.7 | 0.3 | 0.7 | 0 | 0 | 0 |
| | bankssts | 7.4 | 2.5 | 25 | 19.7 | 51.9 | 44.2 | 0.7 | 0.3 | 0.7 | 0.7 | 0 | 0 |
| | precuneus | 2.5 | 4.3 | 15.9 | 18.9 | 37.7 | 41.6 | 0 | 0 | 0.7 | 0.7 | 0 | 0 |
| | superiorparietal | 4.3 | 4.3 | 14.4 | 14.4 | 41.6 | 36.4 | 0.3 | 0.3 | 0.7 | 1.4 | 0 | 0 |
| | supramarginal | 3.7 | 2.5 | 15.2 | 15.2 | 35.1 | 36.4 | 0.7 | 0.7 | 0 | 0 | 0 | 0 |
| | transversetemporal | 1.8 | 4.3 | 9.8 | 9.8 | 19.5 | 22.1 | 0.3 | 1.3 | 2.1 | 2.1 | 0 | 0 |
| $V_2$ | parsopercularis | 1.8 | 1.2 | 11.4 | 10.6 | 26 | 23.4 | 0.7 | 0.7 | 0 | 1.4 | 0 | 0 |
| | parsorbitalis | 1.2 | 1.2 | 6.8 | 7.6 | 16.9 | 19.5 | 0 | 0 | 0 | 0 | 0 | 0 |
| | parstriangularis | 1.2 | 1.2 | 6.8 | 5.3 | 20.8 | 18.2 | 0.3 | 0.7 | 0 | 0 | 0 | 0 |
| | frontalpole | 1.2 | 0.6 | 3.8 | 4.5 | 15.6 | 16.9 | 0 | 0.3 | 0 | 0 | 0 | 0 |
| | caudalmiddlefrontal | 3.1 | 2.5 | 15.9 | 19.7 | 37.7 | 37.7 | 0.3 | 0.7 | 0.7 | 2.1 | 0 | 0 |
| | lateralorbitofrontal | 0.6 | 0.6 | 9.8 | 9.1 | 22.1 | 18.2 | 0 | 0 | 0.7 | 0.7 | 0 | 0 |
| | medialorbitofrontal | 1.2 | 0.6 | 4.5 | 8.3 | 14.3 | 18.2 | 0.3 | 0 | 0 | 1.4 | 0 | 0 |
| | rostralmiddlefrontal | 1.8 | 3.7 | 12.1 | 15.2 | 22.1 | 23.4 | 0 | 0 | 0 | 0 | 0 | 0 |
| | superiorfrontal | 1.8 | 2.5 | 8.3 | 9.8 | 23.4 | 23.4 | 0 | 0 | 0.7 | 0.7 | 0 | 0 |
| VI | pericalcarine | 0.6 | 0.6 | 6.8 | 7.6 | 23.4 | 15.6 | 0.3 | 0 | 0.7 | 0.7 | 0 | 0 |
| | cuneus | 1.8 | 1.2 | 12.9 | 11.4 | 31.2 | 28.6 | 0 | 0 | 0.7 | 0 | 0 | 0 |
| | paracentral | 3.1 | 1.8 | 6.1 | 4.5 | 18.2 | 13 | 0.7 | 0.3 | 2.8 | 1.4 | 0 | 0 |
| | postcentral | 1.8 | 2.5 | 4.5 | 3 | 16.9 | 20.8 | 0.3 | 1 | 0 | 0 | 0 | 0 |
| | precentral | 1.2 | 1.8 | 8.3 | 6.1 | 20.8 | 26 | 0.3 | 0.7 | 0.7 | 1.4 | 0 | 0 |

CU = Cognitively unimpaired, MCI = Mild cognitive impairment, AD = Alzheimer's disease, LH = Left hemisphere, RH = Right hemisphere, Aβ = Amyloid. $V_2$ represents Braak V region in the frontal lobe while $V_1$ regroups the remaining Braak V regions. An asterisk next to the region name indicate that this region is part of the temporal meta region of interest (Jack et al., 2017)

**Supplementary Table 3 – Regional tau abnormality progression across regions of interest**

| | | CU Aβ+ (n = 90) | | MCI Aβ+ (n = 66) | | AD Aβ+ (n = 39) | | CU Aβ- (n = 96) | | MCI Aβ- (n = 40) | | AD Aβ- (n = 10) | |
|---|---|---|---|---|---|---|---|---|---|---|---|---|---|
| **Braak stage** | **Region** | **LH** | **RH** | **LH** | **RH** | **LH** | **RH** | **LH** | **RH** | **LH** | **RH** | **LH** | **RH** |
| I | entorhinal * | 8.9 | 14.4 | 12.1 | 4.5 | 10.3 | 5.1 | 3.1 | 1.0 | 5.0 | 2.5 | 0.0 | 0.0 |
| III | amygdala * | 3.3 | 6.7 | 10.6 | 9.1 | 2.6 | 0.0 | 1.0 | 1.0 | 2.5 | 2.5 | 10.0 | 0.0 |
| | fusiform * | 4.4 | 4.4 | 16.7 | 12.1 | 2.6 | 10.3 | 2.1 | 1.0 | 5.0 | 2.5 | 0.0 | 0.0 |
| | parahippocampal * | 7.8 | 7.8 | 6.1 | 13.6 | 5.1 | 5.1 | 1.0 | 3.1 | 2.5 | 5.0 | 10.0 | 0.0 |
| | lingual | 2.2 | 1.1 | 10.6 | 9.1 | 2.6 | 2.6 | 2.1 | 1.0 | 0.0 | 0.0 | 0.0 | 0.0 |
| IV | temporalpole | 6.7 | 5.6 | 12.1 | 16.7 | 12.8 | 15.4 | 1.0 | 1.0 | 2.5 | 0.0 | 10.0 | 0.0 |
| | inferiortemporal * | 4.4 | 4.4 | 12.1 | 13.6 | 2.6 | 7.7 | 2.1 | 2.1 | 2.5 | 7.5 | 0.0 | 0.0 |
| | middletemporal * | 5.6 | 6.7 | 12.1 | 12.1 | 5.1 | 2.6 | 1.0 | 1.0 | 2.5 | 0.0 | 0.0 | 0.0 |
| | isthmuscingulate | 4.4 | 2.2 | 10.6 | 4.5 | 7.7 | 5.1 | 1.0 | 2.1 | 0.0 | 0.0 | 0.0 | 0.0 |
| | caudalanteriorcingulate | 4.4 | 0.0 | 6.1 | 4.5 | 2.6 | 0.0 | 2.1 | 1.0 | 0.0 | 0.0 | 0.0 | 0.0 |
| | insula | 4.4 | 6.7 | 7.6 | 6.1 | 12.8 | 7.7 | 2.1 | 3.1 | 2.5 | 0.0 | 0.0 | 0.0 |
| | posteriorcingulate | 4.4 | 3.3 | 10.6 | 7.6 | 7.7 | 7.7 | 1.0 | 2.1 | 0.0 | 0.0 | 0.0 | 0.0 |
| | rostralanteriorcingulate | 2.2 | 1.1 | 3.0 | 3.0 | 5.1 | 0.0 | 2.1 | 1.0 | 0.0 | 0.0 | 0.0 | 0.0 |
| $V_1$ | lateraloccipital | 2.2 | 2.2 | 12.1 | 9.1 | 5.1 | 7.7 | 1.0 | 1.0 | 2.5 | 2.5 | 0.0 | 0.0 |
| | inferiorparietal | 1.1 | 4.4 | 15.2 | 13.6 | 0.0 | 2.6 | 1.0 | 2.1 | 0.0 | 0.0 | 0.0 | 0.0 |
| | superiortemporal | 4.4 | 7.8 | 7.6 | 6.1 | 5.1 | 10.3 | 1.0 | 2.1 | 2.5 | 0.0 | 0.0 | 0.0 |
| | bankssts | 1.1 | 5.6 | 6.1 | 9.1 | 0.0 | 2.6 | 1.0 | 1.0 | 2.5 | 0.0 | 0.0 | 0.0 |
| | precuneus | 2.2 | 2.2 | 12.1 | 12.1 | 5.1 | 0.0 | 2.1 | 1.0 | 0.0 | 0.0 | 0.0 | 0.0 |
| | superiorparietal | 3.3 | 3.3 | 10.6 | 12.1 | 2.6 | 7.7 | 1.0 | 1.0 | 0.0 | 0.0 | 0.0 | 0.0 |
| | supramarginal | 4.4 | 4.4 | 10.6 | 12.1 | 12.8 | 2.6 | 1.0 | 1.0 | 0.0 | 0.0 | 0.0 | 0.0 |
| | transversetemporal | 4.4 | 1.1 | 3.0 | 9.1 | 10.3 | 2.6 | 2.1 | 3.1 | 0.0 | 0.0 | 0.0 | 0.0 |
| $V_2$ | parsopercularis | 4.4 | 2.2 | 7.6 | 12.1 | 5.1 | 5.1 | 2.1 | 2.1 | 0.0 | 0.0 | 0.0 | 0.0 |
| | parsorbitalis | 2.2 | 1.1 | 3.0 | 0.0 | 2.6 | 0.0 | 2.1 | 3.1 | 0.0 | 0.0 | 0.0 | 0.0 |
| | parstriangularis | 2.2 | 1.1 | 1.5 | 9.1 | 5.1 | 0.0 | 2.1 | 2.1 | 0.0 | 0.0 | 0.0 | 0.0 |
| | frontalpole | 2.2 | 2.2 | 1.5 | 4.5 | 2.6 | 0.0 | 1.0 | 0.0 | 0.0 | 0.0 | 0.0 | 0.0 |
| | caudalmiddlefrontal | 3.3 | 7.8 | 7.6 | 4.5 | 2.6 | 2.6 | 2.1 | 0.0 | 0.0 | 0.0 | 0.0 | 0.0 |
| | lateralorbitofrontal | 3.3 | 1.1 | 7.6 | 7.6 | 0.0 | 0.0 | 2.1 | 2.1 | 0.0 | 0.0 | 0.0 | 0.0 |
| | medialorbitofrontal | 2.2 | 2.2 | 7.6 | 7.6 | 7.7 | 2.6 | 1.0 | 4.2 | 0.0 | 0.0 | 0.0 | 0.0 |
| | rostralmiddlefrontal | 4.4 | 5.6 | 4.5 | 3.0 | 7.7 | 2.6 | 2.1 | 2.1 | 0.0 | 0.0 | 0.0 | 0.0 |
| | superiorfrontal | 3.3 | 4.4 | 10.6 | 7.6 | 5.1 | 0.0 | 3.1 | 2.1 | 0.0 | 0.0 | 0.0 | 0.0 |
| VI | pericalcarine | 1.1 | 1.1 | 0.0 | 1.5 | 2.6 | 5.1 | 3.1 | 1.0 | 0.0 | 0.0 | 0.0 | 0.0 |
| | cuneus | 0.0 | 3.3 | 4.5 | 4.5 | 5.1 | 2.6 | 3.1 | 1.0 | 0.0 | 0.0 | 0.0 | 0.0 |
| | paracentral | 1.1 | 2.2 | 9.1 | 6.1 | 12.8 | 7.7 | 3.1 | 1.0 | 0.0 | 0.0 | 0.0 | 0.0 |
| | postcentral | 0.0 | 2.2 | 7.6 | 1.5 | 5.1 | 7.7 | 1.0 | 1.0 | 0.0 | 0.0 | 0.0 | 0.0 |
| | precentral | 2.2 | 4.4 | 7.6 | 7.6 | 2.6 | 5.1 | 2.1 | 1.0 | 0.0 | 0.0 | 0.0 | 0.0 |

Numbers are in percentages. CU = Cognitively unimpaired, MCI = Mild cognitive impairment, AD = Alzheimer's disease, LH = Left hemisphere, RH = Right hemisphere, Aβ = Amyloid. $V_2$ represents Braak V region in the frontal lobe while $V_1$ regroups the remaining Braak V regions. An asterisk next to the region name indicate that this region is part of the temporal meta region of interest (Jack et al., 2017)
